# Leveraging pleiotropy to improve genetic risk prediction across diseases

**DOI:** 10.1101/2025.06.16.25329688

**Authors:** Jiaqi Hu, Geyu Zhou, Hongyu Zhao, Andrew T. DeWan

**Affiliations:** Department of Chronic Disease Epidemiology, Yale School of Public Health, New Haven, CT 06510, USA; Department of Biological Sciences, Purdue University, West Lafayette, Indiana 47907, USA; Department of Statistics, Purdue University, West Lafayette, Indiana 47907, USA.; Department of Biostatistics, Yale School of Public Health, New Haven, CT 06510, USA; Program of Computational Biology and Bioinformatics, Yale University, New Haven, CT, USA

**Keywords:** polygenic score (PGS), pleiotropy, risk prediction

## Abstract

**Background:** Polygenic scores (PGSs) have shown promise in predicting disease risk, but their predictive accuracy remains limited for many complex diseases. Leveraging the shared genetic architecture among correlated traits may improve prediction performance.

**Methods:** We developed a flexible framework for constructing multi-trait PGSs by integrating candidate PGSs (N=2,651) derived from publicly available GWAS summary statistics (N=51)—using single-trait, MTAG-all, and MTAG-pairwise approaches. Multi-trait PGS models were trained using Elastic Net regression in the UK Biobank (N = 307,230 individuals) and validated in both an internal set of UKB individuals (N = 39,122) and an external, All of Us (N = 116,394), cohort. We further evaluated the utility of multi-trait PGSs in risk prediction with non-genetic factors, interactions, and genetic subgroup identification.

**Results:** Multi-trait PGSs significantly improved risk prediction for eight diseases, with AUC gains ranging from 1.56% to 5.45% compared to optimal single-GWAS PGSs. Selected scores mainly consisted of genetically correlated phenotypes. Multi-trait PGSs further enhanced predictive performance and stratification when integrated with non-genetic factors. Significant interactions were identified between multi-trait PGS for peripheral artery disease (PAD) and modifiable risk factors such as smoking and waist-to-hip ratio (WHR). A clustering analysis uncovered genetically distinct subgroups with meaningful phenotypic variation, including a chronic kidney disease (CKD) subgroup enriched for diabetes- and obesity-related traits.

**Conclusion:** Our multi-trait PGS framework improves disease prediction by capturing cross-trait genetic effects and enables personalized risk assessment through integration with non-genetic exposures, interactions, and subgroup identification. This approach offers a scalable and generalizable tool for advancing precision medicine.

## Introduction

Genome-wide association studies (GWASs) have identified thousands of single-nucleotide polymorphisms (SNPs) associated with various complex traits and diseases^13^. Building on these findings, polygenic scores (PGSs)–which quantify genetic predisposition by aggregating the effects of associated SNPs as a weighted sum of effect alleles–have emerged as a promising tool for improving disease risk prediction and informing preventive strategies. Despite their potential, the predictive performance of current PGSs remains modest for many traits and diseases. This limitation is largely due to the substantial sample sizes required to accurately estimate the small effect sizes of numerous SNPs that together explain trait heritability. To address this challenge, researchers have explored the integration of genetic information across multiple genetically correlated traits or diseases. By leveraging the shared genetic architecture among related phenotypes, multi-trait approaches can enhance the accuracy and generalizability of PGSs.

Methodologies for integrating genetic effects across multiple traits or diseases can be categorized into three main approaches. The first approach involves modeling cross-phenotype genetic effects jointly through specialized statistical algorithms, such as PleioPred and mtPGS. These methods typically require prior assumptions about the genetic structure and cannot be combined with widely used single-trait PGS methods.

The second approach applies meta-analysis to combine summary statistics from multiple GWASs, followed by conventional single-trait PGS methods to derive polygenic weights. An example of this strategy is shaPRS, which accounts for variant-level heterogeneity across GWASs by incorporating this information into a weighted meta-analysis framework. Despite its utility, shaPRS has notable limitations: it currently supports only two input GWASs per analysis and does not account for linkage disequilibrium (LD) when evaluating variant-level heterogeneity.

The third and most widely adopted approach involves a linear or non-linear combination of PGSs derived from multiple traits or diseases. This framework has significantly improved predictive performance, particularly for phenotypes with lower heritability or strong genetic correlations with other traits. The observed gains are likely explained by the identification of individuals with both the target condition and genetically related comorbidities. Crucially, the effectiveness of this approach depends on constructing a diverse and relevant candidate PGS pool encompassing a broad range of genetically correlated phenotypes. To generate such a pool, some studies rely on domain knowledge to select traits or diseases known to be associated with the target disease^10^. While informative, this method is inherently subjective and often tailored to specific disease contexts. Alternatively, a more agnostic strategy involves curating a comprehensive PGS library from publicly available resources, such as the PGS Catalog.

However, this approach requires scrutiny to avoid sample overlap and inconsistencies in population ancestry between the reference studies and the target cohort.

In this study, we developed a novel framework for constructing multi-trait PGSs by combining the second and third approaches. Our method was designed to maintain compatibility with existing single-trait PGS pipelines. Candidate phenotypes were selected through a data-driven strategy, wherein we curated a pool of 51 complex traits and diseases with publicly available, large-scale GWAS summary statistics. Notably, GWASs involving the UK Biobank (UKB) cohort were excluded to avoid sample overlap in downstream analyses. Across-all and pairwise meta-analyses were conducted, and PGS weights were inferred through a single-trait PGS method, SDPR. The final multi-trait PGS models were trained via Elastic Net (EN) models, and the prediction performance was evaluated. Furthermore, we highlight three applications of our framework: (1) enhancing risk prediction of non-genetic risk models, (2) offering individualized preventive strategies via interactions with modifiable factors, and (3) providing personalized therapeutic guidance through genetic subgroup assignment.

Together, our approach provides a scalable and adaptable strategy for leveraging cross-phenotype genetic architecture to improve precision medicine.

## Methods

### Overview of methods

The methodology is illustrated in Figure 1. We began by preparing GWAS summary statistics from three sources: (1) original GWAS summary statistics (excluding UK Biobank [UKB]) for 51 selected phenotypes ("single-trait"); (2) meta-analysis combining all 51 phenotypes simultaneously (“MTAG-all”); and (3) pairwise meta-analyses across the same phenotypes (“MTAG-pairwise”). We then calculated PGS weights from each GWAS source using the single-trait method SDPR (Figure 1A). The resulting PGSs constructed from individual GWASs were hereafter referred to as single-GWAS PGSs.

**Figure 1.**
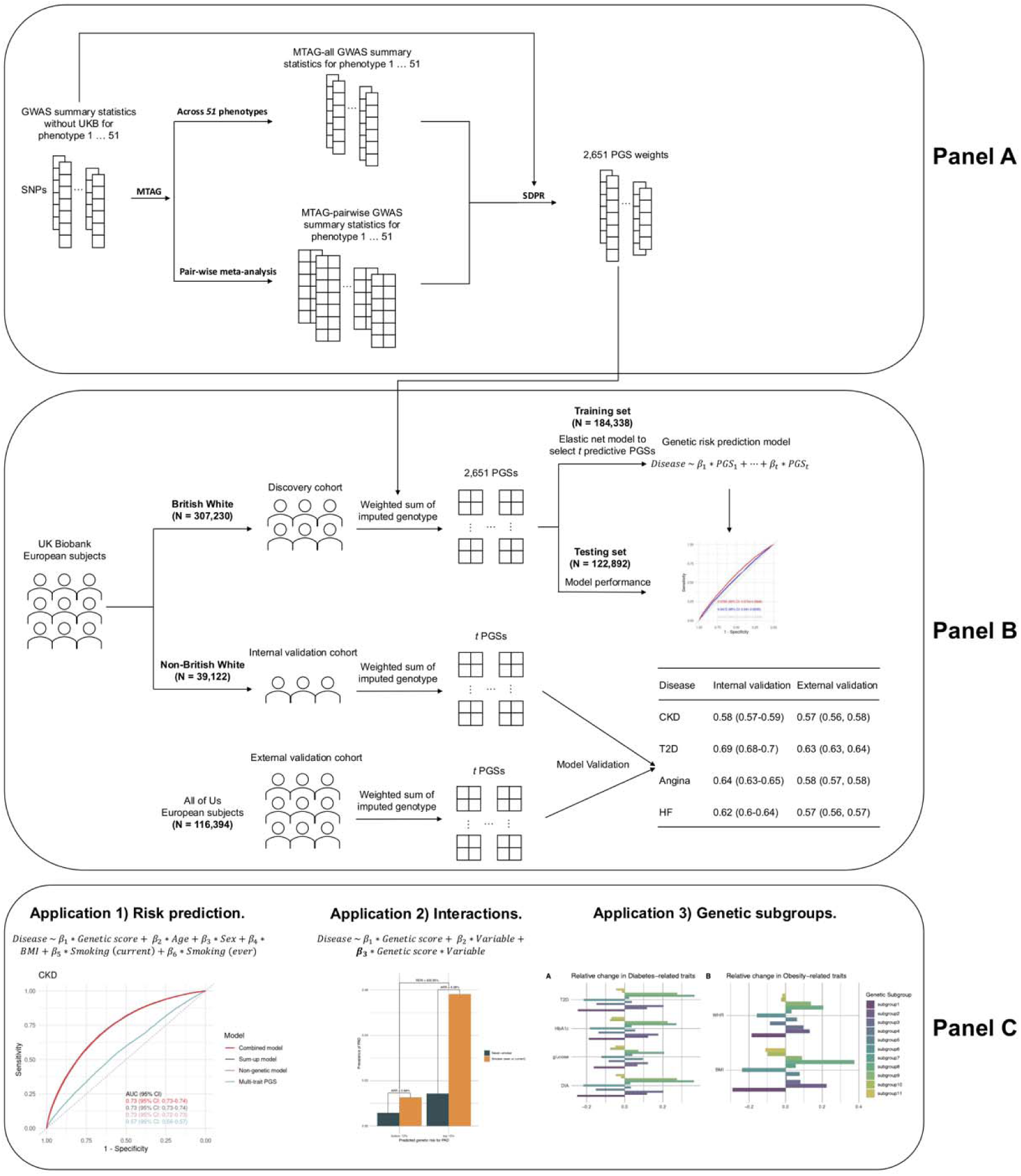
Overview of the multi-trait PGS framework and study design. The overall analytical pipeline consists of three main components: (A) derivation of 2,651 candidate PGSs from 51 GWAS summary statistics using three approaches— single-trait GWASs, MTAG-all, and MTAG-pairwise; (B) training, testing, and validation of multi-trait PGS models in European ancestry individuals from the UKB, with British White subjects divided into training (N = 184,338) and testing (N = 122,892) sets, and Non-British White subjects (N = 39,122) used as an internal validation cohort. External validation was performed in European ancestry individuals from the All of Us cohort (N = 116,394); (C) downstream applications of the multi-trait PGS models, including: (1) integration with non-genetic risk factors for enhanced risk prediction, (2) analysis of interactions with variables of interest, and (3) identification of genetically distinct subgroups. Model performance was evaluated using metrics such as AUC, calibration, ORs, and disease prevalence across risk quantiles.

Next, genetic risk models of multi-trait PGSs were developed and validated using European-ancestry participants from the UKB cohort (Figure 1B). Lastly, three applications of these multi-trait PGSs were considered: (1) integration with non-genetic risk factors for disease prediction, (2) interactions between genetic risk and other risk factors, and (3) identification of genetic subgroups (Figure1C).

### Development of PGSs

Publicly available, large-scale GWAS summary statistics for 51 phenotypes were downloaded (listed in Supplemental Table 1), which were grouped into 14 phenotype clusters based on domain knowledge. All selected GWASs explicitly excluded participants from the UK Biobank (UKB) cohort to avoid sample overlap. We excluded indels and duplicated SNPs from all summary statistics and retained only HapMap3 SNPs present in the UKB genotype data to ensure high-quality, well-imputed variants for downstream analyses.

Three strategies were employed to generate GWAS summary statistics for PGS inference: (1) single-trait: original GWAS summary statistics for each phenotype were directly utilized without modification; (2) MTAG-all: Summary statistics for all 51 phenotypes were jointly meta-analyzed using MTAG, generating 51 combined meta-analysis outputs. MTAG accounts explicitly for sample overlap across summary statistics; (3) Pairwise meta-analyses across the 51 phenotypes were conducted using MTAG, resulting in 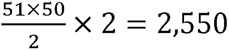 outputs. The latter two strategies (MTAG-all and MTAG-pairwise) explicitly leverage genetic correlations between traits, thus representing multi-trait approaches. In total, these approaches yielded 51+51+2,550=2,652 sets of prepared GWAS summary statistics.

Polygenic score weights were subsequently inferred from these prepared summary statistics using SDPR, a computationally efficient method that does not require parameter tuning. An external LD reference panel comprising 503 European subjects from the 1000 Genomes Project Phase III was used. Finally, single-GWAS PGSs for study subjects were computed using PLINK v1.90.

### Study subjects

Participants from the UKB, enrolled at ages 40 to 69, were included in this analysis. Genotyping was performed using either the UK BiLEVE array or the UK Biobank Axiom array, with 733,332 autosomal variants shared between the two arrays. High-quality genotype data imputed to reference panels (imputation R^2^ greater than 0.8 and minor allele frequency [MAF] greater than 0.001) was used. We restricted the analysis to 346,352 unrelated subjects with genetically confirmed European ancestry, dividing them into two main groups: British White subjects (discovery cohort, N=307,230) and Non-British White subjects (internal validation cohort, N=39,122) (Figure 1B). The discovery cohort was further randomly partitioned into training (60%) and testing (40%) subsets.

Multi-trait PGS models were established using the training subset, with prediction performance evaluated in the testing subset and subsequently validated using the internal validation cohort as well as an external validation dataset (All of Us cohort).

The 51 target phenotypes included in this study were defined within the UKB cohort using corresponding clinical variables available in the UKB database or a combination of self-reported diagnoses and hospital records. Specifically, direct matches for 15 phenotypes were identified using available UKB Field IDs (Supplemental Table 2), while 26 diseases were defined based on either self-reported diagnoses or relevant hospital diagnostic codes (listed in Supplemental Table 2). Alzheimer’s disease (AD) was uniquely defined using a previously validated quantitative proxy derived from parental history of AD and parents’ current age, consistent with the approach described by Jansen et al. (2019). The remaining 10 phenotypes were unavailable in the UKB dataset and thus not assessed directly.

### External validation in All of Us

To evaluate the generalizability of the developed multi-trait PGS models, we conducted external validation using data from the All of Us Researcher Workbench (version 7), which includes both genotype and phenotype information curated by the All of Us Research Program. SNPs included in the multi-trait PGSs were extracted from the All of Us whole-genome sequencing (WGS) data released under the Access-Controlled Aggregate Dataset Format (ACAF), ensuring consistency with the genomic coordinates and variant annotations used in model training. The analysis was restricted to unrelated participants of genetically inferred European ancestry, as determined by the auxiliary ancestry assignments file provided by All of Us. To ensure consistency in phenotype definitions and minimize potential confounding, individuals with non-binary sex assigned at birth were excluded from the odds ratio (OR) analysis. Phenotypes in the All of Us cohort were defined using structured variables and curated clinical concepts available through the Researcher Workbench, including condition occurrence data from electronic health records (EHRs) and self-reported surveys. Individuals were classified as cases if they were included in the corresponding curated patient cohort for each phenotype, based on standardized condition codes. These included self-reported survey data, electronic health records (EHR), and derived clinical conditions available in the curated datasets. The multi-trait PGSs developed in the UKB training dataset were applied to the All of Us genotype data to calculate risk scores with the shared SNPs, and the predictive performance was evaluated using the same modeling framework as in the UKB validation phase. This step provided an independent assessment of model robustness across biobank populations.

### Model derivation and analysis

A total of 2,651 single-GWAS PGSs were computed for each subject, excluding the MTAG-all PGS for fibrinogen due to the absence of valid SNPs with non-zero effect sizes. First, we assessed their prediction accuracy for target phenotypes via R^2^ for quantitative phenotypes and area under the receiver operating characteristic curve (AUC) for binary ones. Then, Elastic Net (EN) regression was used to construct multi-trait PGSs for 25 binary disease outcomes in the training dataset, with model tuning performed via 10-fold cross-validation to identify the optimal lambda minimizing prediction error.

Model performance was evaluated in the testing dataset across four dimensions: (1) discrimination: assessed by AUC; (2) calibration: evaluated using calibration plots and the Hosmer–Lemeshow goodness-of-fit test; (3) relative risk: estimated through ORs adjusted for age at recruitment, sex, and the top 10 genetic principal components (PCs); (4) absolute risk: examined via disease prevalence across quantiles of the predicted score. The final models were validated in both the UKB internal validation set and the All of Us external cohort using the same evaluation metrics. In the All of Us cohort, covariates included sex and the top 10 PCs.

To interpret the multi-trait PGS models, we quantified feature importance (FI) as the absolute value of the EN coefficients. PGSs were grouped into 14 phenotype clusters based on domain knowledge (Supplemental Table 1). The cluster FIs were calculated as the mean FI within each group, and contributions were computed as the proportion of cluster FIs.

To explore the hypothesis that the superior performance of multi-trait PGSs can be explained by the ability to capture comorbidity with genetically correlated diseases^5^, we introduced a discordance score, defined as the difference between standardized multi-trait and single-trait PGS predictions. Among disease cases, we tested whether higher discordance scores were associated with an increased burden of related conditions. A positive association would support the hypothesis that multi-trait models enhance prediction by identifying individuals with comorbidities. Of note, to test the robustness of our definition, we conducted a sensitivity analysis with the discordance score defined as the differences between the predicted values of multi-trait PGSs without standardization.

### Application 1. Enhancing risk prediction of non-genetic models

To evaluate whether adding multi-trait PGSs can enhance models with non-genetic risk factors, we trained models that integrated genetic and non-genetic risk factors using two strategies: (1) Sum-up strategy: Separate models for genetic and non-genetic components were trained in the training set. The predicted scores were then summed linearly in the testing and validation sets; and (2) Combined strategy: A single model incorporating all 2,651 PGSs and non-genetic variables was trained directly in the training set and evaluated in the testing and validation cohorts. Non-genetic predictors included age at recruitment, sex, body mass index (BMI), and smoking status. Although a minimal set was used here for simplicity, the framework is readily extendable to include additional covariates.

To enhance statistical power for evaluation, the testing and internal validation samples were combined. Model performance was assessed using the same metrics applied to the genetic models: AUC for discrimination, calibration plots and Hosmer–Lemeshow tests for calibration, ORs adjusted for covariates, and disease prevalence across score quantiles. The AUCs were compared via Delong tests.

### Application 2. Identifying interactions

One important application of PGSs is to explore interactions between aggregated genetic risk and non-genetic variables, such as lifestyle factors and clinical biomarkers. By employing multi-trait PGSs—which provide more accurate estimates of genetic risk—we may uncover a greater number of meaningful interactions. Therefore, we hypothesized that individuals with higher multi-trait PGSs may experience greater adverse effects from specific exposures. To test this, we selected 23 variables encompassing baseline measurements, biomarkers, smoking behaviors, and medication use and evaluated their interactions with multi-trait PGSs across diseases of interest. Models were adjusted for age at recruitment, sex, and the top 10 genetic PCs when applicable.

Due to the composite nature of multi-trait PGSs, correlations with the exposure variables may introduce multicollinearity, potentially inflating interaction effects^22^. To test the robustness of identified interactions to multicollinearity, we conducted a sensitivity analysis: each variable of interest was regressed out of the multi-trait PGS using linear regression, and interactions were re-tested using the resulting residuals. Interactions with consistent direction and magnitude were considered robust to multicollinearity.

Beyond statistical interaction, we assessed absolute interaction using the Relative Excess Risk due to Interaction (RERI). Specifically, we compared absolute risk (AR)— defined as disease prevalence—between exposed and unexposed individuals in the top and bottom deciles of the multi-trait PGS distribution. The absolute risk reduction (ARR) due to non-exposure was calculated separately in each risk group, and RERI was defined as the difference in ARRs between the high- and low-risk groups, normalized by the baseline absolute risk in unexposed individuals in the bottom decile. Thus, RERI quantified the proportion of excess cases attributable to the interaction between genetic and non-genetic factors. The statistical significance of the RERI was tested using the delta method^23^.

### Application 3. Classifying subjects into genetic subgroups

Emerging evidence suggests that genetic heterogeneity among patients with the same disease may be linked to phenotypic variation and differential treatment response^24^.

Leveraging the PGSs selected in our multi-trait prediction models, we constructed individual-level genetic profiles for disease cases and identified subgroups based on shared genetic architecture.

Specifically, we calculated weighted PGSs for each subject in the testing and internal validation cohorts using the coefficients from the trained EN models. Hierarchical clustering was then applied to the resulting genetic profile matrix. The optimal number of clusters was determined using the elbow method based on the total within-cluster sum of squares.

To characterize genetic heterogeneity, we compared the contributions of each PGS group—defined by phenotype clusters (Supplemental Table 1)—across the identified subgroups. Phenotypic differences between subgroups and the remaining cases were assessed through association testing and summarized by relative change, defined as the difference in means (for continuous variables) or proportions (for binary variables) between the subgroup and others, normalized by the standard deviation of the variable.

This approach enabled us to uncover genetically defined subpopulations within common diseases, offering insights into disease mechanisms and potential opportunities for stratified medicine.

### Statistical analysis

Unless otherwise specified, multiple comparisons were controlled using the Bonferroni correction. All statistical analyses were conducted using R (version 4.2).

## Results

### Model development and assessment

The demographic characteristics of the study population are summarized in Table 1. Using 2,651 candidate single-GWAS PGSs derived from 51 single-trait GWASs, 50 MTAG-all GWASs, and 2,550 MTAG-pairwise GWASs, we first assessed their prediction performance for corresponding phenotypes in the UK Biobank testing dataset. For continuous traits, prediction accuracy (R^2^) for single-trait PGSs ranged from 0.17% (95% CI 0.08%-0.25%) for age starting smoking (AgeSmk) to 9.35% (95% CI 9.06%- 9.66%) for HDL, and the R^2^ for MTAG-pairwise PGSs ranged from 0.28% (95% CI 0.16%-0.38%) for AgeSmk to 10.53% (95% CI 10.21%-10.86%) for HDL. However, MTAG-all PGSs achieved R^2^ values below 0.3% across all traits (Supplemental Table 3).

**Table 1.** Demographic characteristics.

| Variable | All subjects<br>(N=346,352) | CKD<br>(N=24,664) | T2D<br>(N=25,292) | Angina<br>(N=24,069) | HF<br>(N=6,094) | Asthma<br>(N=48,741) | PAD<br>(N=6,381) | CAD<br>(N=23,563) | IS<br>(N=16,210) |
| --- | --- | --- | --- | --- | --- | --- | --- | --- | --- |
| Age at recruitment | 56.72±8.03* | 61.41±6.57 | 60.10±6.90 | 61.37±6.26 | 62.10±6.14 | 56.40±8.18 | 61.37±6.61 | 61.19±6.41 | 61.46±6.41 |
| Sex (Male) | 159,216<br>(45.97%)<sup>#</sup> | 13,567<br>(55.01%) | 15,353<br>(60.70%) | 15,541<br>(64.57%) | 3,942<br>(64.69%) | 2,0780<br>(42.63%) | 4,196<br>(65.76%) | 17,599<br>(74.69%) | 11,186<br>(69.01%) |
| BMI | 27.38±4.76 | 29.35±5.53 | 31.70±5.74 | 29.23±5.04 | 30.23±6.06 | 28.22±5.34 | 28.56±5.30 | 28.92±4.75 | 28.76±4.96 |
| Smoking |  |  |  |  |  |  |  |  |  |
| Never | 187,441<br>(54.12%) | 10,749<br>(43.58%) | 10,581<br>(41.84%) | 9,727 (40.41%) | 2,343<br>(38.45%) | 25,707<br>(52.74%) | 1,743<br>(27.32%) | 9,082 (38.54%) | 6,188 (38.17%) |
| Previous | 122,653<br>(35.41%) | 10,512<br>(42.62%) | 11,360<br>(44.92%) | 11,260<br>(46.78%) | 2,832<br>(46.47%) | 17,900<br>(36.72%) | 3,016<br>(47.27%) | 11,118<br>(47.18%) | 7,618 (47.00%) |
| Current | 36,258 (10.47%) | 3,403 (13.80%) | 3,351 (13.25%) | 3,082 (12.80%) | 919 (15.08%) | 5,134 (10.53%) | 1,622<br>(25.42%) | 3,363 (14.27%) | 2,404 (14.83%) |
| Alcohol intake frequency |  |  |  |  |  |  |  |  |  |
| Daily or almost daily | 73,848 (21.32%) | 4,906 (19.89%) | 4,070 (16.09%) | 4,742 (19.70%) | 1,291<br>(21.18%) | 9,878 (20.27%) | 1,577<br>(24.71%) | 5,063 (21.49%) | 3,546 (21.88%) |
| Three or four times a week | 82,886 (23.93%) | 4,479 (18.16%) | 4,291 (16.97%) | 4,923 (20.45%) | 1,125<br>(18.46%) | 10,836<br>(22.23%) | 1,164<br>(18.24%) | 5,148 (21.85%) | 3,392 (20.93%) |
| Once or twice a week | 91,056 (26.29%) | 6,035 (24.47%) | 6,269 (24.79%) | 6,059 (25.17%) | 1,412<br>(23.17%) | 12,241<br>(25.11%) | 1,461<br>(22.90%) | 5,961 (25.30%) | 3,967 (24.47%) |
| One to three times a month | 38,493 (11.11%) | 2,725 (11.05%) | 3,148 (12.45%) | 2,528 (10.50%) | 631 (10.35%) | 5,649 (11.59%) | 601 (9.42%) | 2,433 (10.33%) | 1,636 (10.09%) |
| Special occasions only | 37,053 (10.70%) | 3,742 (15.17%) | 4,407 (17.42%) | 3,308 (13.74%) | 881 (14.46%) | 6,100 (12.52%) | 879 (13.78%) | 2,817 (11.96%) | 2,026 (12.50%) |
| Never | 23,016 (6.65%) | 2,777 (11.26%) | 3,107 (12.28%) | 2,509 (10.42%) | 754 (12.37%) | 4,037 (8.28%) | 699 (10.95%) | 2,141 (9.09%) | 1,643 (10.14%) |
| SBP | 139.73±19.02 | 144.59±20.11 | 144.63±18.59 | 142.81±19.23 | 143.32±20.88 | 139.40±18.64 | 144.71±20.30 | 143.42±19.53 | 142.18±19.85 |
| DBP | 82.15±10.32 | 82.58±10.90 | 82.95±10.53 | 81.11±10.67 | 81.52±11.53 | 82.27±10.24 | 80.99±10.81 | 81.72±10.80 | 81.32±10.93 |
| HDL | 1.45±0.36 | 1.35±0.36 | 1.23±0.30 | 1.29±0.32 | 1.32±0.36 | 1.44±0.36 | 1.32±0.36 | 1.27±0.31 | 1.30±0.34 |
| LDL | 3.56±0.85 | 3.34±0.92 | 3.09±0.92 | 3.18±0.94 | 3.20±0.90 | 3.54±0.85 | 3.21±0.92 | 3.22±0.96 | 3.08±0.90 |
| TG | 1.74±1.00 | 1.97±1.11 | 2.25±1.26 | 1.98±1.12 | 1.95±1.13 | 1.76±1.01 | 1.98±1.16 | 2.00±1.13 | 1.89±1.07 |
| TC | 5.71±1.12 | 5.38±1.24 | 5.00±1.23 | 5.14±1.25 | 5.18±1.23 | 5.68±1.13 | 5.20±1.24 | 5.17±1.28 | 5.02±1.21 |

CKD: chronic kidney disease; T2D: type 2 diabetes; HF: heart failure; PAD: peripheral artery disease; CAD: coronary artery disease; IS: ischemic stroke.
\*: mean±standard deviation; #: number (proportion).

For binary outcomes, single-trait PGSs yielded AUCs ranging from 0.5 (95% CI 0.49-0.51) for ischemic stroke (IS) to 0.72 (95% CI 0.69-0.75) for schizophrenia (SCZ), and the MTAG-pairwise PGSs improved AUCs ranging from 0.54 (95% CI 0.51-0.56) for ovarian cancer to 0.72 (95% CI 0.69-0.75) for SCZ. The MTAG-all PGSs performed poorly with AUCs around 0.5 for most outcomes. Notably, MTAG-pairwise PGSs outperformed single-trait PGSs for most phenotypes, with statistically significant improvements (P < 0.05) observed in 27 traits. Exceptions included alcohol intake frequency, breast cancer, and prostate cancer, where single-trait PGSs performed better. Interestingly, for most of the 27 phenotypes with improved performance, the best MTAG-pairwise PGSs were derived from the meta-analysis of the target phenotype and a closely related condition. For example, the top-performing PGS for peripheral artery disease (PAD) resulted from a pairwise meta-analysis of PAD and coronary artery disease (CAD). These findings highlight the importance of SNP coverage in PGS construction and demonstrate that incorporating genetically correlated traits via pairwise meta-analysis can substantially enhance prediction accuracy.

We next developed multi-trait PGS prediction models for 25 binary disease outcomes using the training dataset. PGSs with non-zero coefficients selected in the final EN models for these eight diseases were listed in Supplemental Table 4. The discriminative prediction, measured by AUC, was compared across the single-trait, the best MTAG- pairwise, and multi-trait PGSs, with results summarized in Table 2. Using the DeLong test, we identified eight diseases–chronic kidney disease (CKD), type 2 diabetes (T2D), angina, heart failure (HF), asthma, PAD, CAD, and ischemic stroke (IS)–for which the multi-trait PGS significantly outperformed the optimal single-trait PGS (P < 0.05). The improvement in AUC ranged from 1.56% for CAD to 5.45% for CKD. ROC curves comparing single-trait, best MTAG-pairwise, and multi-trait PGSs were shown in Figure 2. Calibration assessments indicated that the multi-trait models were well-calibrated across all eight diseases (Supplemental Figure 1). In terms of relative risk, the multi-trait PGS consistently yielded higher ORs compared to both single-trait and MTAG-pairwise PGSs (Supplemental Table 5). For example, the multi-trait PGS for CKD showed an OR of 1.36 (95% CI: 1.33–1.39), representing a 20.35% increase over the single-trait PGS (OR = 1.13, 95% CI: 1.10–1.15) and a 14.29% increase over the best MTAG-pairwise PGS (OR = 1.19, 95% CI: 1.16–1.22). Absolute risk was evaluated by disease prevalence across polygenic score quantiles (Supplemental Figure 2). Multi-trait PGSs consistently showed the greatest risk stratification. For instance, the prevalence of HF was 3.79% among individuals in the top 5% of the multi-trait PGS distribution, compared to 3.06% for MTAG-pairwise and 2.82% for single-trait. Overall, across multiple performance metrics, the multi-trait PGS models demonstrated superior predictive accuracy over both single-trait and MTAG-pairwise approaches in eight diseases.

**Figure 2.**
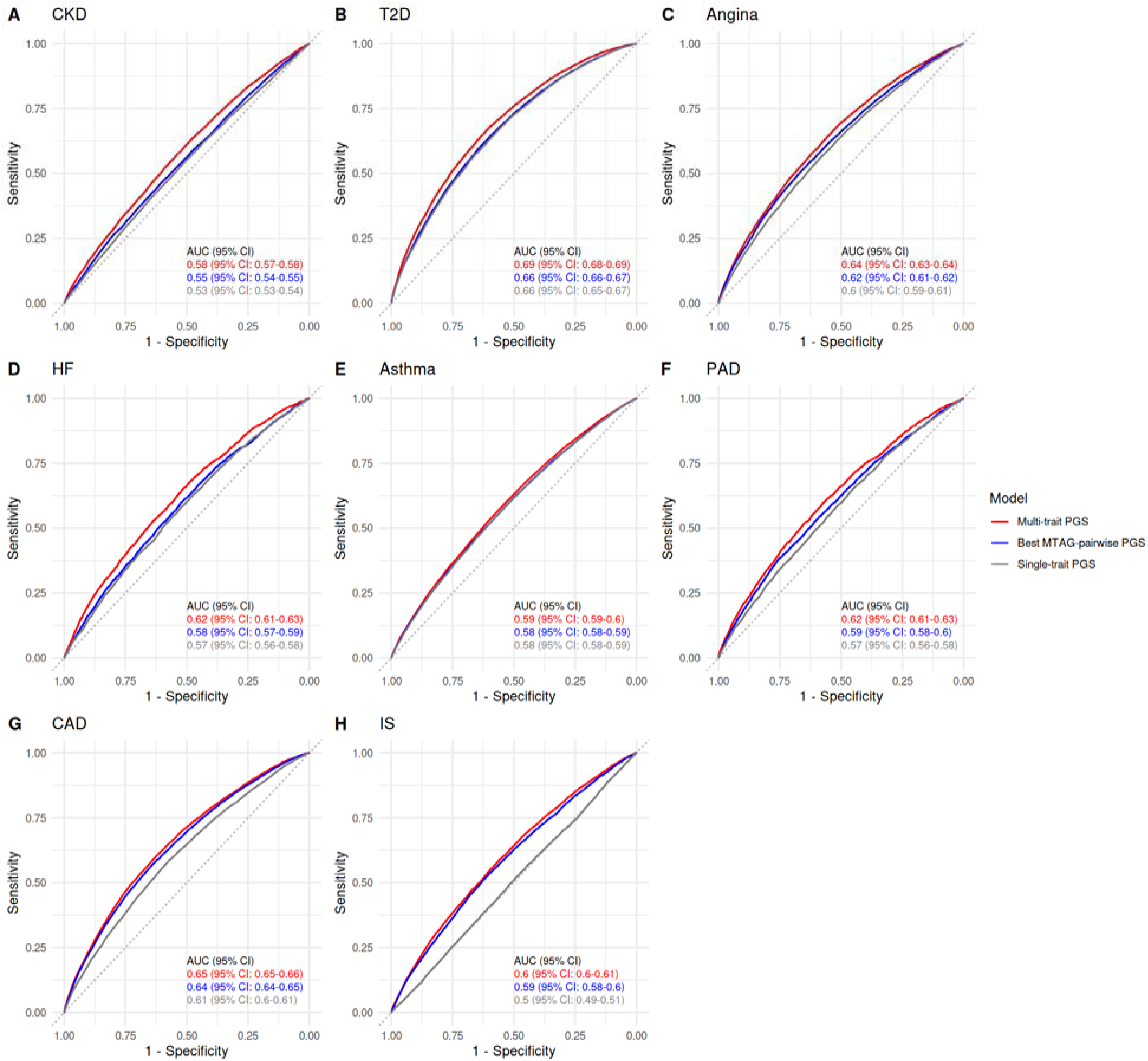
Comparison of PGS performance across three models for eight diseases. The ROC curves for disease prediction using three PGS models: single-trait PGS (gray), the best MTAG-pairwise PGS (blue), and multi-trait PGS (red). Results were shown for eight diseases with significant improvement in prediction using the multi-trait PGS: (A) CKD, (B) T2D, (C) angina, (D) HF, (E) asthma, (F) PAD, (G) CAD, and (H) IS. the AUC and 95% confidence intervals were displayed in each panel. In all eight diseases, the multi-trait PGS outperformed both the single-trait and best MTAG-pairwise models in terms of discriminative accuracy.

**Table 2.** Comparison of AUCs of the best single-GWAS and multi-trait PGSs.

| Disease | Best single-GWAS PGS | Multi-trait PGS | P-value |
| --- | --- | --- | --- |
| CKD | 0.55 (95% CI: 0.54-0.55) | 0.58 (95% CI: 0.57-0.58) | 2.42E-12 |
| T2D | 0.66 (95% CI: 0.66-0.67) | 0.69 (95% CI: 0.68-0.69) | 1.30E-09 |
| Angina | 0.63 (95% CI: 0.61-0.62) | 0.64 (95% CI: 0.63-0.64) | 2.87E-05 |
| HF | 0.58 (95% CI: 0.57-0.59) | 0.62 (95% CI: 0.61-0.63) | 4.83E-05 |
| Asthma | 0.58 (95% CI: 0.58-0.59) | 0.59 (95% CI: 0.59-0.6) | 3.72E-03 |
| PAD | 0.59 (95% CI: 0.58-0.6) | 0.62 (95% CI: 0.61-0.63) | 4.53E-03 |
| CAD | 0.64 (95% CI: 0.64-0.65) | 0.65 (95% CI: 0.65-0.66) | 9.62E-03 |
| IS | 0.59 (95% CI: 0.58-0.6) | 0.6 (95% CI: 0.6-0.61) | 1.27E-02 |
| ADHD | 0.65 (95% CI: 0.55-0.74) | 0.48 (95% CI: 0.37-0.59) | 2.35E-02 |
| RA | 0.57 (95% CI: 0.56-0.58) | 0.59 (95% CI: 0.58-0.6) | 7.87E-02 |
| Ovarian cancer | 0.54 (95% CI: 0.51-0.56) | 0.51 (95% CI: 0.49-0.54) | 1.58E-01 |
| ASD | 0.68 (95% CI: 0.59-0.77) | 0.57 (95% CI: 0.42-0.72) | 2.27E-01 |
| Lung cancer | 0.6 (95% CI: 0.58-0.61) | 0.61 (95% CI: 0.59-0.62) | 2.34E-01 |
| Osteoporosis | 0.57 (95% CI: 0.56-0.58) | 0.57 (95% CI: 0.57-0.58) | 2.75E-01 |
| IBD | 0.64 (95% CI: 0.63-0.66) | 0.65 (95% CI: 0.64-0.66) | 4.52E-01 |
| AF | 0.64 (95% CI: 0.63-0.65) | 0.65 (95% CI: 0.64-0.65) | 4.99E-01 |
| PSC | 0.56 (95% CI: 0.53-0.58) | 0.55 (95% CI: 0.52-0.58) | 6.90E-01 |
| SLE | 0.66 (95% CI: 0.62-0.69) | 0.65 (95% CI: 0.61-0.69) | 7.94E-01 |
| PAH | 0.58 (95% CI: 0.54-0.62) | 0.57 (95% CI: 0.53-0.61) | 8.04E-01 |
| BIP | 0.67 (95% CI: 0.64-0.7) | 0.67 (95% CI: 0.65-0.7) | 8.07E-01 |
| MDD | 0.58 (95% CI: 0.57-0.58) | 0.58 (95% CI: 0.57-0.58) | 8.36E-01 |
| SCZ | 0.72 (95% CI: 0.69-0.75) | 0.72 (95% CI: 0.69-0.75) | 8.87E-01 |
| PBC | 0.81 (95% CI: 0.76-0.85) | 0.81 (95% CI: 0.76-0.85) | 9.86E-01 |
| Breast cancer | 0.65 (95% CI: 0.65-0.66) | 0.65 (95% CI: 0.65-0.66) | 9.92E-01 |
| Prostate cancer | 0.69 (95% CI: 0.68-0.7) | 0.69 (95% CI: 0.68-0.7) | 9.92E-01 |

PGS: polygenic score; CKD: chronic kidney disease; T2D: type 2 diabetes; HF: heart failure; PAD: peripheral artery disease; CAD: coronary artery disease; IS: ischemic stroke; ADHD: attention deficit hyperactivity disorder; RA: rheumatoid arthritis; ASD: autism spectrum disorder; IBD: inflammatory bowel disease; AF: atrial fibrillation; PSC: primary sclerosing cholangitis; SLE: Systemic Lupus Erythematosus; PAH: pulmonary arterial hypertension; BIP: bipolar disorder; MDD: major depression disorder; SCZ: schizophrenia; PBC: primary biliary cholangitis.

The multi-trait PGS models for the eight diseases were further validated in two independent cohorts: non-British White subjects from the UK Biobank (N = 39,122) and unrelated individuals of European ancestry from the All of Us cohort (N = 116,394). In the internal validation cohort, the multi-trait PGSs demonstrated comparable AUCs to those observed in the UKB testing set. In the external validation cohort, AUCs were modestly reduced for seven diseases except for CKD, with decreasing rates ranging from 3.33% for IS to 12.12% for CAD (Supplemental Table 6). The calibration remained strong across diseases (Supplemental Figure 3). Relative and absolute risk patterns were consistent with the testing set results, though the magnitude of ORs was attenuated in the All of Us cohort (Supplemental Table 7 and Supplemental Figure 4).

For example, the OR for CAD was 1.91 (95% CI: 1.83-2) in the internal validation while decreased to 1.37 (95% CI: 1.35-1.4) in the external validation. These findings suggest that the differences across cohorts may have an impact on prediction accuracy of the multi-trait PGSs.

### Model analysis

The relative contributions of each PGS group to the multi-trait models across the eight diseases are presented in Figure 3. For all eight diseases, the PGS group corresponding to the target disease or closely related phenotypes ranked among the top one or two contributors out of the 14 phenotype groups. For example, PGSs related to atherosclerotic traits accounted for 17.89% of the multi-trait PGS for CAD, the highest among all contributing groups. In addition to disease-specific contributions, genetically correlated phenotype groups also played a substantial role. For instance, body measurement–related PGSs contributed 24.81% to the multi-trait PGS for T2D, consistent with strong and significant genetic correlations between the T2D and traits such as body mass index (BMI) and waist-to-hip ratio (WHR) (Supplemental Figure 5).

**Figure 3.**
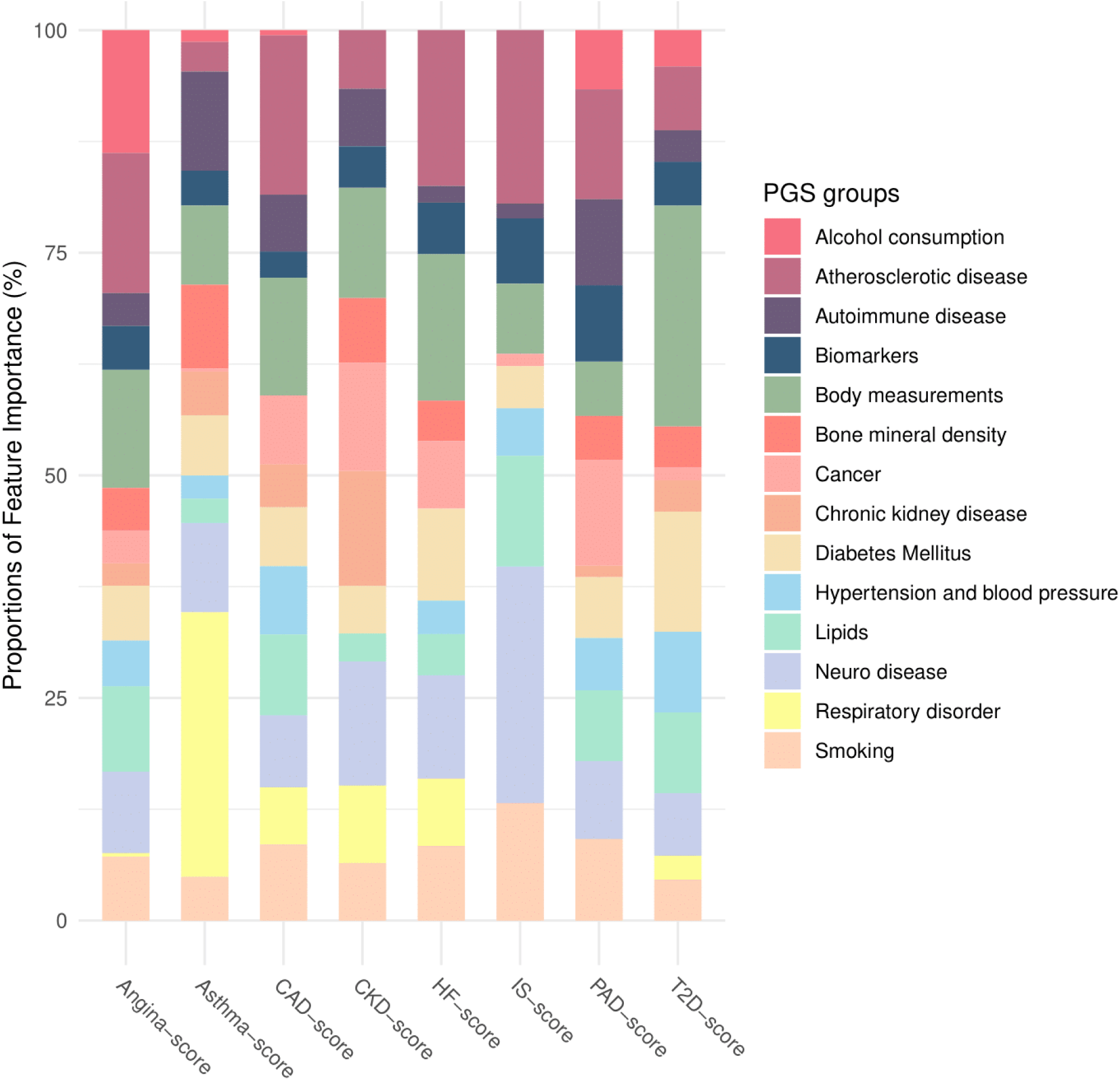
Contributions of PGS phenotype groups to multi-trait PGS models across eight diseases. Bar plots showing the proportion of total feature importance contributed by each of 14 PGS phenotype groups in the multi-trait PGS models for eight diseases. Feature importance was calculated as the absolute value of model coefficients from the EN regression, and values were averaged within each phenotype group. The results highlight the dominant contributions of disease-relevant groups (e.g., body measurements and diabetes for T2D, neuropsychiatric traits for IS), as well as genetically correlated trait groups, reflecting the pleiotropic structure leveraged by the multi-trait PGS framework.

Similarly, neuropsychiatric traits—primarily attention-deficit/hyperactivity disorder (ADHD) and major depressive disorder (MDD)—contributed 26.56% to the multi-trait PGS for IS. These findings underscore the value of incorporating genetically correlated traits in PGS construction, supporting the hypothesis that leveraging shared genetic architecture can enhance risk prediction beyond single-trait models.

Association analyses of the discordance score—defined as the difference between standardized multi-trait and single-trait PGS predictions—revealed consistent patterns across diseases, highlighting significant links with high-burden comorbidities (Figure 4). For instance, among asthma patients, the discordance score was strongly associated with obesity-related traits, such as BMI (β = 1.83, P = 3.85 × 10^-145^). Similarly, in T2D cases, higher discordance scores were associated with increased cardiovascular risk, including a significant association with CAD (β = 0.30, P = 3.51 × 10^-12^). Sensitivity analyses using alternative definitions of the discordance score yielded similar results, confirming the robustness of these associations (Supplemental Figure 6). Together, these findings support the hypothesis that the improved predictive performance of multi-trait PGSs arises, at least in part, from their ability to capture individuals with underlying comorbidities, thereby enhancing clinical relevance.

**Figure 4.**
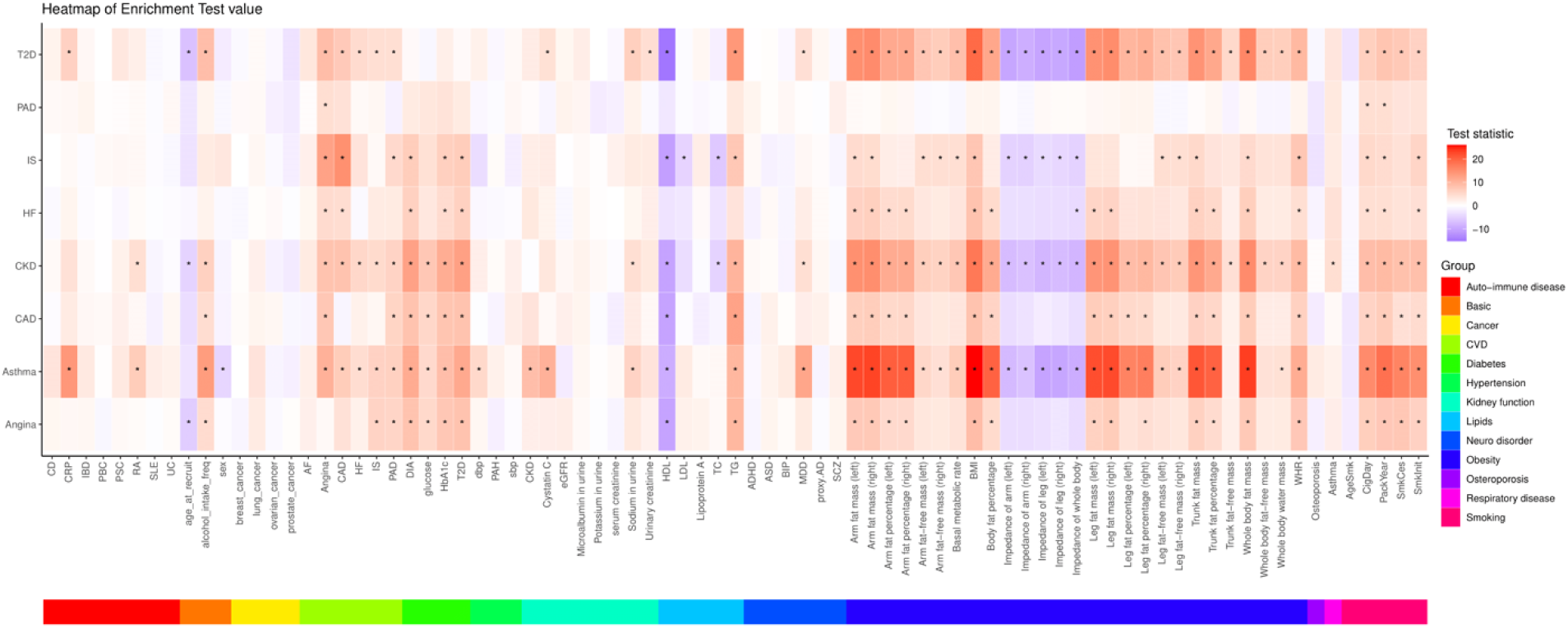
Phenotypic associations with discordance scores across eight diseases. Heatmap showing standardized association statistics (z-scores) between the discordance score and 86 phenotypes among cases of eight diseases. The discordance score was defined as the difference between standardized multi-trait PGS predictions and single-trait PGS predictions. Rows represent disease-specific analyses, and columns represent phenotypes grouped into 13 clinical or biological categories. Asterisks indicate phenotypes with statistically significant associations after Bonferroni correction. Notably, high discordance scores were associated with comorbid traits such as obesity, cardiovascular biomarkers, and neuropsychiatric disorders, supporting the hypothesis that multi-trait PGSs capture individuals with greater disease burden due to comorbidity.

### Risk prediction along with non-genetic factors

We evaluated the integrated models combining multi-trait PGSs with non-genetic risk factors–age at recruitment, sex, BMI, and smoking–using two strategies: the sum-up model, which linearly combined independently trained genetic and non-genetic models, and the combined model, which trained a unified model incorporating all predictors simultaneously. Both integrated models significantly outperformed the non-genetic model regarding discrimination (Figure 5, Supplemental Table 8). For example, across the eight diseases, the average AUC improvement of the combined model over the non-genetic model ranged from 0.63% for CKD to 1.27% for T2D. The combined model also demonstrated superior calibration to the sum-up model, as evidenced by calibration plots and Hosmer–Lemeshow statistics (Supplemental Figure 7). Regarding risk stratification, both integrated models showed substantially higher ORs and disease prevalence in the extreme risk quantiles compared to the non-genetic model (Supplemental Table 9, Supplemental Figure 8). For example, among individuals in the top 5% risk group of asthma, the combined model yielded up to a 3.41-fold increase in disease prevalence over the bottom 10%, compared to a 2.08-fold increase observed with the non-genetic model. No statistically significant differences in ORs or prevalence were observed between the sum-up and combined strategies. These findings demonstrate that multi-trait PGSs can be seamlessly integrated into existing non-genetic risk models, substantially improving disease prediction and enabling better risk stratification in clinical settings.

**Figure 5.**
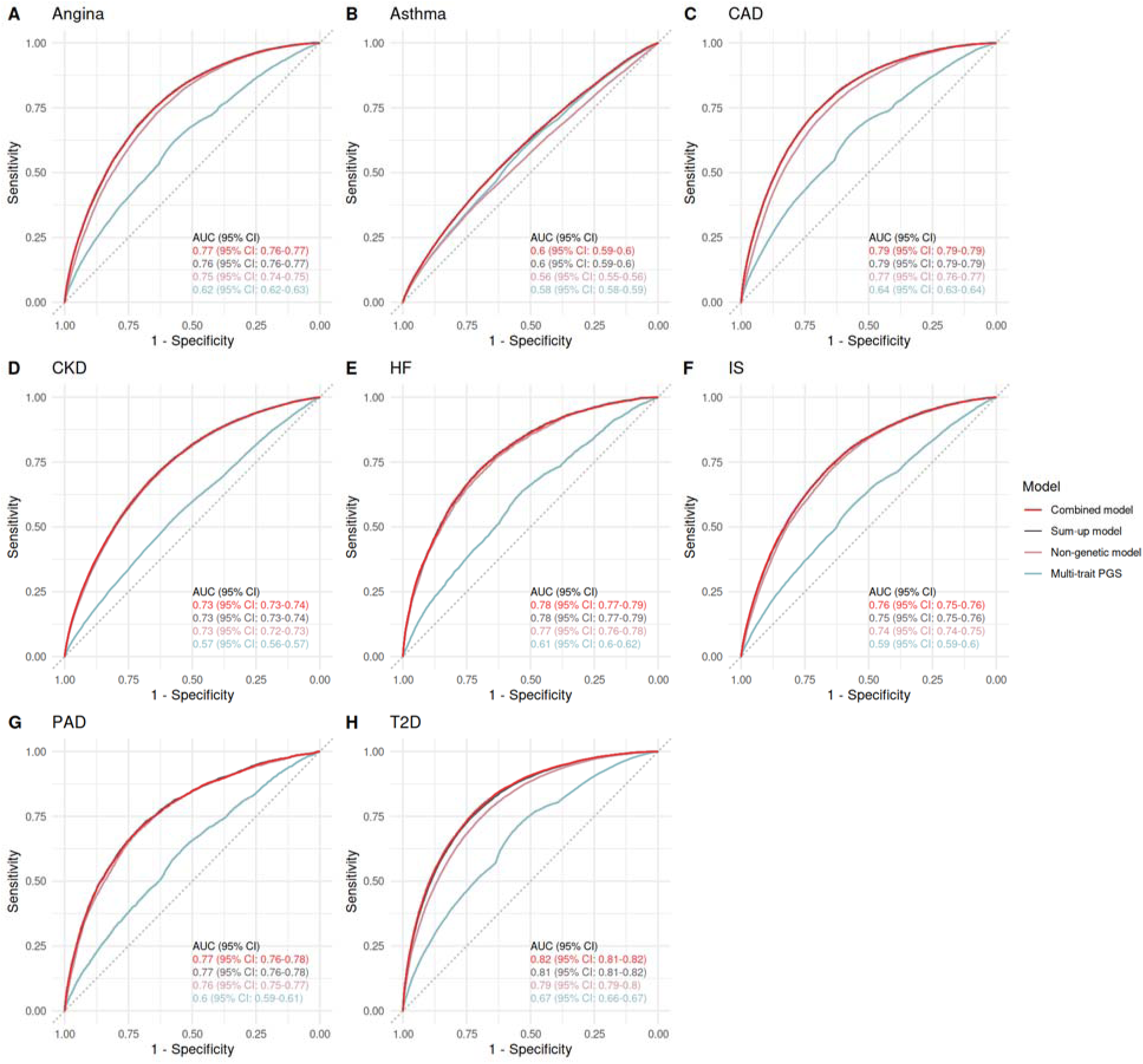
Risk prediction performance of models integrating multi-trait PGSs and non-genetic factors across eight diseases. The ROC curves comparing four risk prediction models: multi-trait PGS (blue), non-genetic model using age, sex, BMI, and smoking status (pink), sum-up model (purple), and combined model (red). AUCs with 95% confidence intervals were displayed within each panel. Across all diseases, the combined model achieved the highest AUC with no significant differences with the sum-up model, indicating improved predictive performance through integration of genetic and clinical risk factors.

### Interactions

We tested interactions between the multi-trait PGSs and 23 non-genetic variables across the eight diseases of interest. After applying Bonferroni correction for multiple testing (significance threshold = 0.05 / (23 × 8) = 2.72 × 10^-4^), 18 PGS–variable pairs exhibited statistically significant interactions (Supplemental Table 10). Notably, the multi-trait PGS for PAD showed significant interactions with SmkInit and WHR. These interactions remained robust after adjusting for multicollinearity. Specifically, the interaction between the PAD PGS and SmkInit yielded a β of 0.17 (P = 6.29 × 10^-5^), and the interaction with WHR produced a β of 0.70 (P = 1.04 × 10^-3^), both of which were nominally significant even after residualizing the PGS to remove correlation with the respective variable.

Furthermore, we analyzed the absolute interactions using ARR and RERI (Figure 6). Both interaction pairs demonstrated strong absolute interaction effects. For SmkInit, the ARR between current or former smokers and never-smokers was 0.68% among individuals in the bottom 10% of the multi-trait PAD PGS. In contrast, the ARR increased to 4.38% in the top 10% risk group, corresponding to a RERI of 635.65% (P = 3.07×10^-10^). This indicates that the excess risk due to smoking was substantially amplified in individuals with high genetic risk. For WHR, individuals were classified as having abdominal obesity based on World Health Organization (WHO) guidelines: WHR ≥ 0.85 for females and ≥ 0.90 for males. Comparing absolute risk between exposed and non-exposed individuals across genetic risk strata, we observed a significantly greater ARR in the top 10% PGS group compared to the bottom 10% group. This corresponded to a RERI of 454.24% (P = 1.56×10^-10^), suggesting a strong synergistic effect between genetic predisposition and abdominal obesity for PAD. These results indicate that a substantial proportion of PAD risk may be attributed to interactions between multi-trait genetic risk and modifiable factors such as smoking and WHR, underscoring their importance in precision prevention strategies.

**Figure 6.**
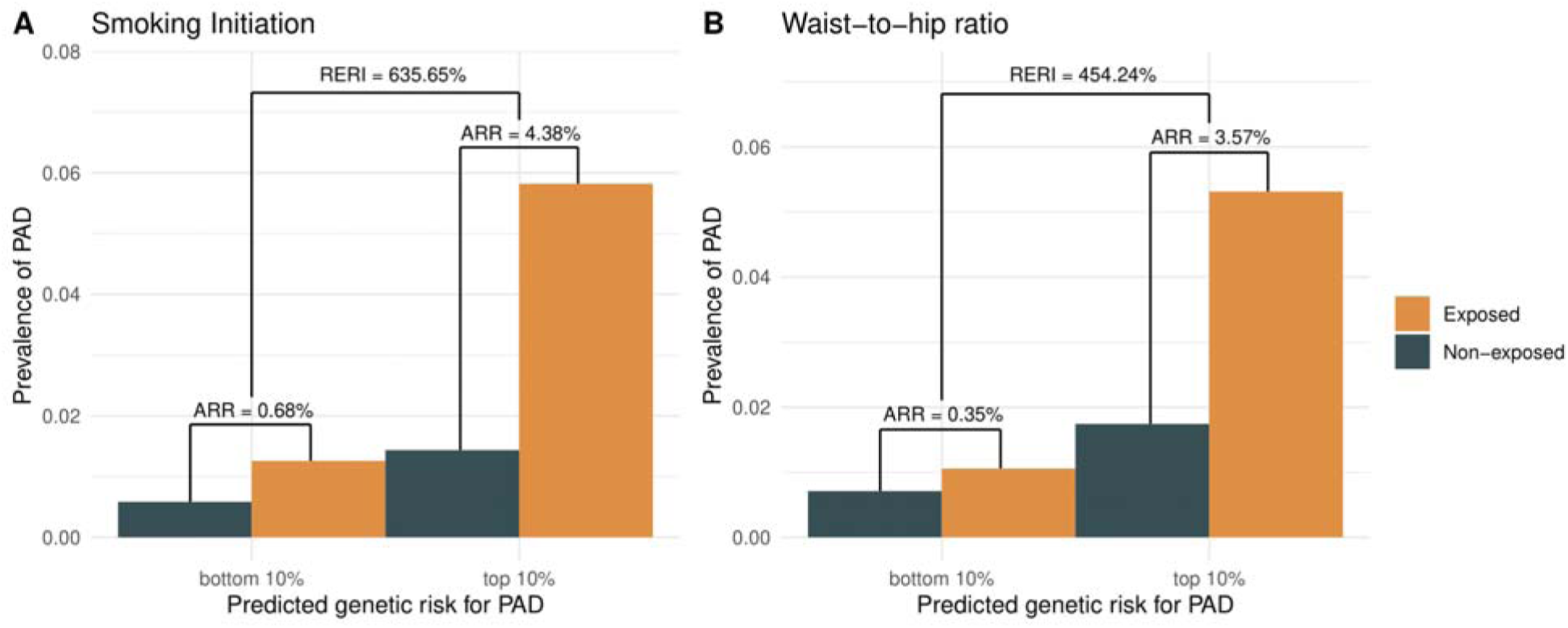
Absolute interaction effects between multi-trait PGS for PAD and smoking and WHR. Absolute risk (AR), reflected by prevalence, of PAD across genetic risk strata and exposure groups for two modifiable risk factors: (A) smoking initiation and (B) WHR. Individuals were stratified into the bottom 10% and top 10% of predicted genetic risk based on the multi-trait PGS for PAD. ARR between exposed and non-exposed individuals was calculated within each risk stratum. The RERI was computed to quantify absolute interaction. Strong absolute interactions were observed for both traits, with higher genetic risk amplifying the impact of exposures to smoking and obesity on PAD prevalence.

### Genetic subgroups

To explore underlying genetic heterogeneity, we identified genetic subgroups within cases of the eight diseases using the selected PGSs from the multi-trait models.

Hierarchical clustering was applied to genetic profiles, and the optimal number of subgroups was determined via the elbow method. The number of subgroups varied by disease, ranging from 9 (e.g., for T2D) to 13 (e.g., for HF) (Supplemental Figure 9). We evaluated subgroup-specific genetic architecture by quantifying the relative contributions of the 14 PGS groups to each cluster. Distinct PGS composition patterns emerged across subgroups within each disease, reflecting substantial heterogeneity in genetic risk sources (Figure 7). For example, in CKD, subgroup 8 displayed markedly elevated contributions from the body measurements and diabetes mellitus PGS groups—accounting for 9.43% and 15.83% of the total feature importance, respectively—compared to mean contributions of 7.71% and 12% across the remaining subgroups (Supplemental Table 11).

**Figure 7.**
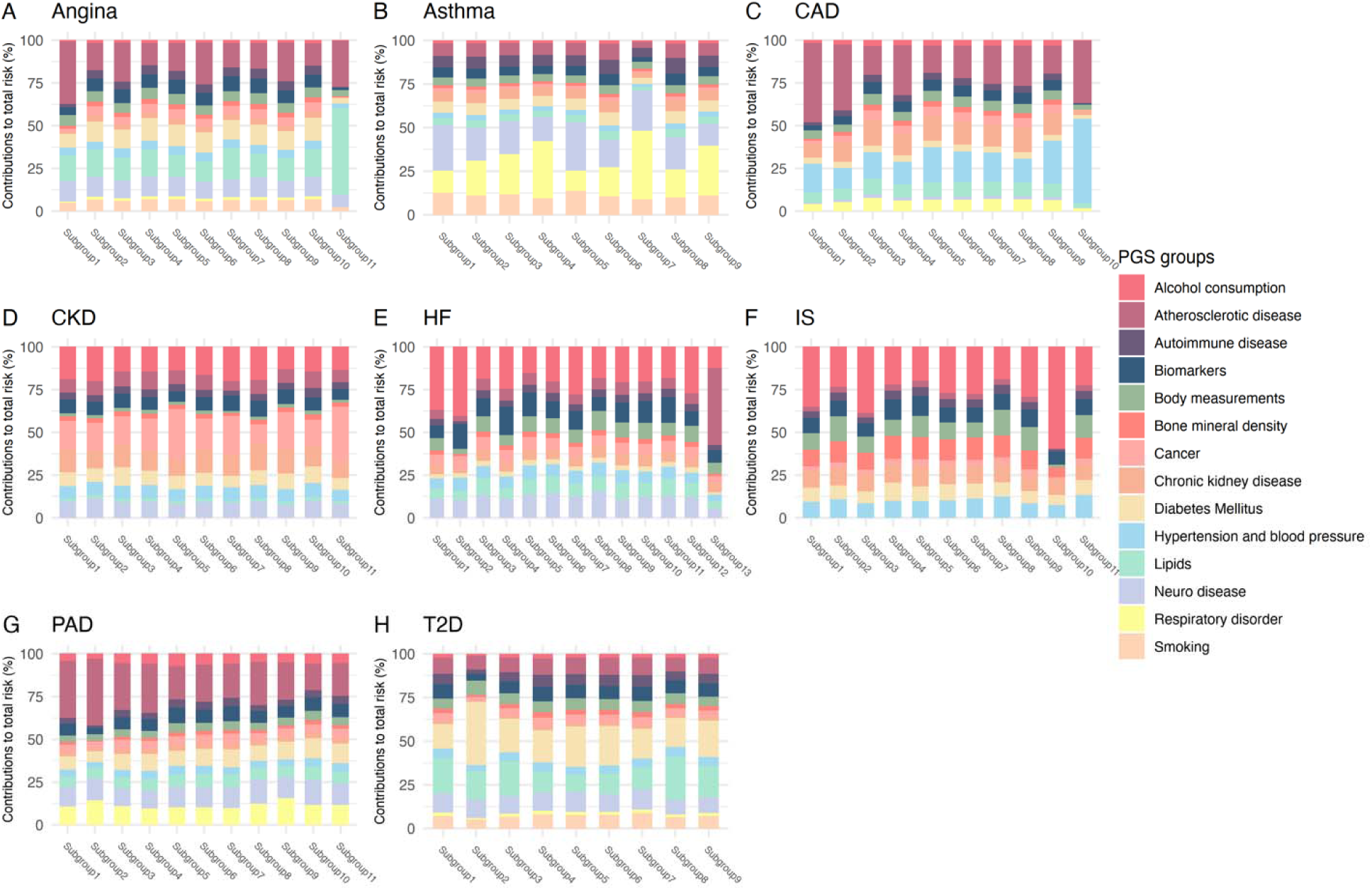
Subgroup-specific contributions of PGS phenotype groups across eight diseases. Stacked bar plots showing the proportional contributions of 14 PGS phenotype groups to the total multi-trait PGS risk within each identified genetic subgroup for eight diseases. Subgroups were defined by hierarchical clustering of disease cases based on their selected PGS profiles. Each bar represents a subgroup, with stacked segments indicating the relative contribution of each phenotype group to the subgroup’s genetic risk. Results reveal distinct subgroup-specific genetic architectures, with some subgroups dominated by contributions from comorbid trait groups, reflecting meaningful heterogeneity in genetic risk composition.

We assessed phenotypic heterogeneity among the genetically defined subgroups using two complementary approaches: (1) comparisons of phenotype distributions between each subgroup and the remaining disease cases; and (2) quantification of relative differences, defined as the standardized difference in means or proportions (Figure 8 and Supplemental Figure 10). Across all eight diseases, we observed clear phenotypic distinctions between genetic subgroups. Notably, in CKD, subgroup 8 exhibited a distinct clinical profile, characterized by a significant difference in T2D distribution with the remaining CKD cases (β = 0.74, P = 2.49×10^-19^). This subgroup also showed elevated glucose (relative difference = 20.81%) and HbA1c levels (relative difference = 27.1%), as well as higher BMI (relative difference = 37.51%) and WHR (relative difference = 20.59%), indicating a strong obesity-related and metabolic burden. These phenotypic patterns closely mirrored the subgroup’s underlying genetic profile, which had heightened contributions from body measurements and diabetes-related PGSs.

**Figure 8.**
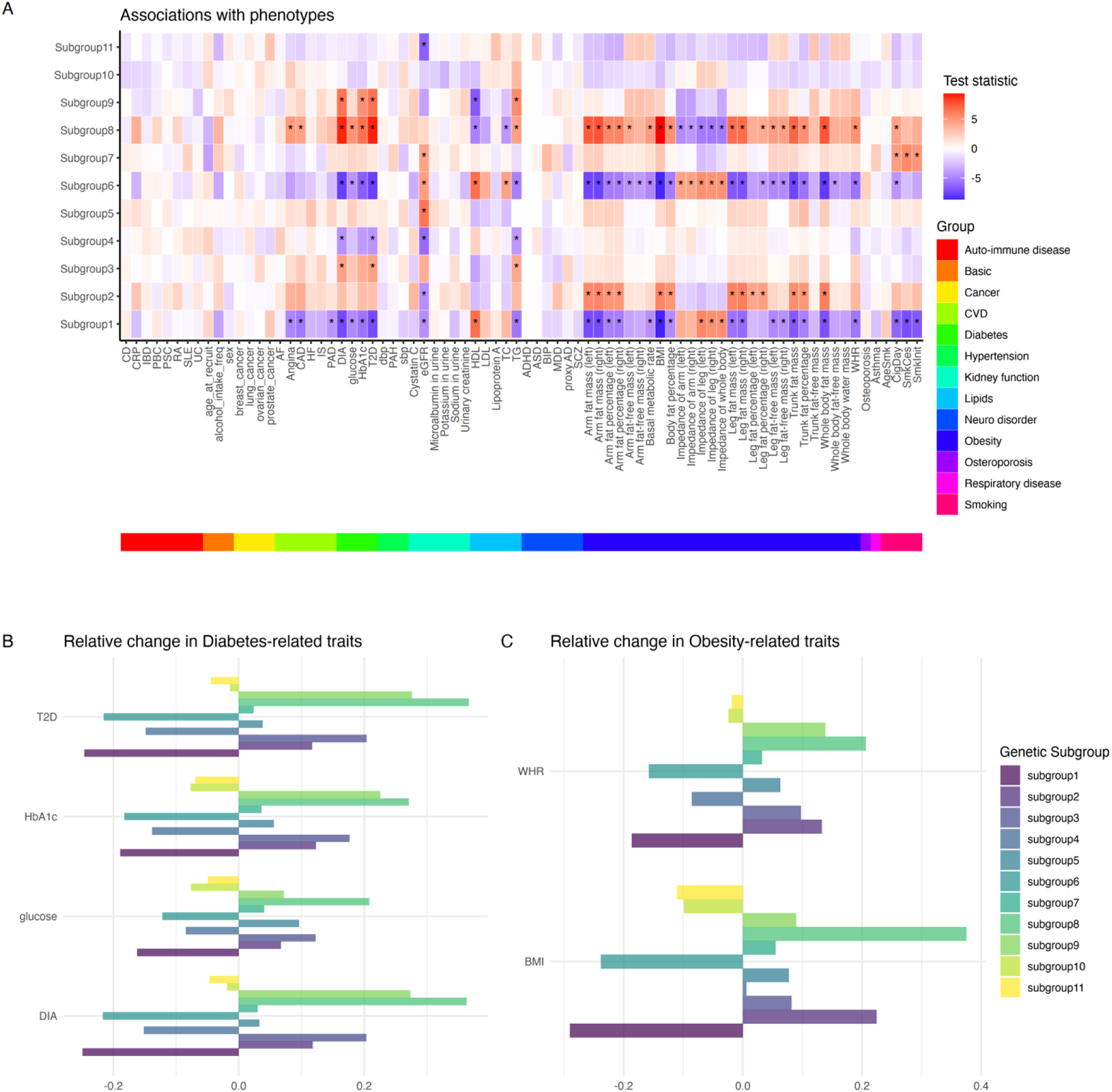
Phenotypic heterogeneity among genetic subgroups in CKD. (A) Heatmap showing statistical associations between CKD genetic subgroups (rows) and 86 clinical and biomarker phenotypes (columns), grouped by biological category. Values represent test statistics (e.g., t-scores), with red indicating positive and blue indicating negative associations. Asterisks denote significant associations after Bonferroni correction. Subgroups exhibit distinct phenotypic profiles. For example, strong associations were found between subgroup 8 and diabetes- and obesity-related phenotypes. These results highlight the heterogeneity in clinical presentations. (B) and (C) Relative changes in diabetes- and obesity-related phenotypes across CKD subgroups. Subgroup 8 showed marked enrichment in both diabetes and obesity traits, consistent with its genetic profile. Together, these results demonstrate that genetically defined subgroups in CKD are associated with distinct clinical characteristics, supporting the potential for stratified risk assessment and personalized management strategies.

These results confirmed that genetic subgroups not only reflect distinct genetic risk profiles but also corresponded to measurable clinical differences, offering potential utility for precision risk stratification and targeted intervention.

## Discussion

We developed multi-trait PGSs by integrating over 2,600 scores derived from 51 GWAS summary statistics and demonstrated significantly improved predictive performance over single-trait PGSs for eight diseases. This enhancement was largely driven by including genetically correlated traits, enabling better identification of individuals with comorbid conditions. Building on these results, we proposed three key applications of multi-trait PGSs: improving risk prediction alongside non-genetic factors, uncovering interactions, and identifying genetically distinct subgroups to support individualized prevention and treatment strategies.

Although a direct comparison with existing multi-trait PGS methods was not feasible due to differences in analytical design, our framework offers several advantages over each of the three major methodological categories. Compared to joint modeling approaches, our method maintains full compatibility with single-trait PGS tools and avoids strong prior assumptions about effect size distributions. While we used SDPR in this study, the framework allows for easy substitution with other PGS inference methods. In contrast to meta-analysis-based approaches, our method accommodates multiple GWAS inputs simultaneously and leverages single-trait PGS methods that account for LD, enabling more accurate modeling of genetic architecture. Furthermore, by incorporating PGSs from three sources–single-trait, MTAG-all, and MTAG-pairwise–we capture a broader spectrum of genetic information from both diseases of interest and disease-relevant traits. Relative to simple PGS-combination strategies, our framework adopts a more rigorous approach to candidate GWAS selection, using only summary statistics that exclude UKB participants and ensuring consistency in ancestry. Additionally, the PGS pool is agnostic to specific target diseases, making it highly scalable and adaptable for future extensions.

By examining the PGSs selected in the multi-trait PGS models across eight diseases, we observed substantial contributions from phenotypes genetically correlated with the target diseases. Several findings aligned with prior research. For instance, PGSs for MDD and ADHD had high FI in the multi-trait PGS for IS, consistent with prior studies highlighting genetic overlap between IS and these neuropsychiatric conditions^25–27^.

These results underscore the role of pleiotropy–where a single genetic variant influences multiple phenotypes–in enhancing polygenic prediction^28^. Incorporating pleiotropic effects enables improved effect size estimation for variants shared across traits.

We also have some interesting observations of pleiotropy. In the case of CKD, the neuropsychiatric PGS group—including ADHD and MDD—contributed the largest proportion of overall feature importance. While epidemiological studies have documented links between impaired kidney function and neuropsychiatric or neurodegenerative disorders^29^, genetic evidence has been limited. One recent study reported potential pleiotropy between kidney function biomarkers and bipolar disorder (BIP) and MDD^30^. Our results provide further support for shared genetic architecture between renal function and neuropsychiatric traits.

Additionally, we noted that some phenotypes included in the prediction models for eight diseases showed no significant genetic correlations with the corresponding disease. For example, the single-trait PGS of prostate cancer was included in the multi-trait PGS for IS (Supplemental Table 4), while no significant genetic correlation was observed between prostate cancer and IS (Supplemental Figure 5). Many of these scores included were derived from the lung cancer GWAS, likely reflecting shared environmental risk through smoking. The other three cancers—breast, ovarian, and prostate cancers—showed little correlations with the eight diseases (Supplemental Figure 5), and their inclusion suggests the presence of local genetic correlations with diseases like PAD. These associations may be obscured in genome-wide genetic correlation estimates due to background noise^31^. Future studies should investigate regional genetic correlations to better characterize these localized pleiotropic effects.

The multi-trait PGSs developed in this study have potential applications in risk prediction and informing individualized prevention and therapeutic strategies. Notably, we identified significant positive interactions between the multi-trait PGS for PAD and two modifiable risk factors: smoking initiation and WHR. Smoking is a well-established and potent risk factor for PAD^32,33^. Our findings suggest that individuals with high genetic risk, as indicated by the multi-trait PGS, may derive greater benefit from smoking cessation efforts, highlighting an opportunity for more targeted clinical interventions. Furthermore, our results reinforce the role of central adiposity—measured by WHR—as a key risk factor for PAD. Prior studies have shown that WHR may be more strongly associated with PAD than other obesity-related measures^33^, and we observed particularly pronounced effects among individuals in the highest genetic risk strata. These interactions support the integration of multi-trait PGSs into precision prevention frameworks, enabling more personalized recommendations for high-risk individuals.

Genetic heterogeneity–reflected in distinct polygenic risk profiles–has been documented across a range of complex diseases^24^. In our study, stratifying CKD patients by their selected PGSs revealed a genetic subgroup marked by a higher prevalence of diabetes and elevated metabolic markers, including glucose levels, WHR, and BMI. These findings are consistent with recent research highlighting shared genetic architecture between CKD and diabetic microvascular complications located in the kidney^34^. The co-occurrence of CKD and diabetes is known to exacerbate disease progression and increase clinical burden, often requiring intensive management of glycemic control, lipid levels, and blood pressure^35^. The subgroup identified in our analysis may, therefore, represent individuals at particularly high risk for diabetic kidney complications. Early identification of such individuals could inform timely preventive strategies and support tailored therapeutic interventions aimed at reducing long-term morbidity.

Several limitations should be acknowledged. First, although candidate phenotypes were selected using a data-driven approach, selection bias may persist due to factors such as disease prevalence and data availability. Notably, most included phenotypes were cardiometabolic, which may partly explain the greater predictive improvement observed for cardiometabolic diseases. However, as more large-scale GWASs become publicly available, expanding the phenotype pool will be straightforward in future work. Second, our analysis included both prevalent and incident cases to maximize sample size, though genetic contributions may differ between the two. For instance, prevalent cases are often enriched for early-onset disease, and polygenic scores are known to have stronger associations with early-onset phenotypes. Validation using incident-only cases will be important in future studies. Third, the current framework may be less suitable for diseases with limited case numbers, such as ADHD, where prediction was less stable due to insufficient statistical power and the large number of candidate PGSs. Fourth, when generalizing the multi-trait PGSs to a cohort independent of the discovery data, the prediction accuracy decreased modestly, suggesting that PGSs might be context-specific, and further studies should be conducted to improve the prediction accuracy of PGSs across contexts. Fifth, while the integration of multi-trait PGSs with non-genetic risk factors showed improved prediction, further calibration may be needed when applying the model in new target populations or clinical settings. Finally, our study was restricted to individuals of European ancestry, and the transferability of the developed PGSs to other ancestral groups remains uncertain. Future research should evaluate the generalizability and adaptation of this framework across diverse populations.

## Conclusion

We proposed a scalable framework for constructing multi-trait PGSs by integrating and selecting relevant predictors from single-trait and meta-analyzed GWAS-derived PGSs. This approach led to significant improvements in risk prediction for eight common diseases, with consistent results observed in both internal and independent external validation cohorts. Analyses of the selected PGSs revealed substantial contributions from genetically correlated phenotypes, highlighting the utility of leveraging cross-trait genetic architecture. Beyond improved prediction, the framework demonstrated utility in identifying interactions with modifiable factors and uncovering genetically distinct subgroups with meaningful phenotypic differences. These findings support the application of multi-trait PGSs in personalized prevention, risk stratification, and targeted therapeutic strategies. Overall, our approach offers a flexible and generalizable tool for advancing precision medicine through multi-phenotype genetic integration.

## Supporting information

References to supplementary materials

## Data Availability

The multi-trait PGSs for eight diseases will be available on the PGS catalog (https://www.pgscatalog.org/) once published. The other data that support the findings of this study are available from the corresponding author upon reasonable request.

## Acknowledgements

The authors would like to thank the researchers and participants of the United Kingdom Biobank. All data was accessed as part of project 32285 from the United Kingdom Biobank. We also thank the Yale Center for Research Computing for using the McCleary High Performance Computing cluster.

## Author contributions

J.H., H.Z., and A.D. contributed to the study conception and design. Data requests and analysis were performed by J.H., H.Z., and A.D. G.Z. helped on the analysis for the All of Us dataset. The first draft of the manuscript was written by J.H. and A.D., and all authors commented on the manuscript. All authors read and approved the final manuscript.

## Ethics approval and consent to participate

This research was conducted using the UK Biobank Resource (application number 32285). The UK Biobank study was conducted under generic approval from the National Health Services’ National Research Ethics Service. The present analyses were conducted in accordance with the Declaration of Helsinki and approved by the Human Investigations Committee at Yale University (2000026836 for UK Biobank). The patients/participants provided their written informed consent to participate in this study.

## Consent of publication

All authors agree to publish.

## Declaration of generative AI and AI-assisted technologies in the writing process

During the preparation of this work the authors used ChatGPT in order to improve writing. After using this tool/service, the authors reviewed and edited the content as needed and take full responsibility for the content of the publication.

