## Supplemental Figures for "Leveraging pleiotropy to improve genetic risk prediction across diseases"

**Supplemental Figure 1. Calibrations of three PGS models across eight diseases.**

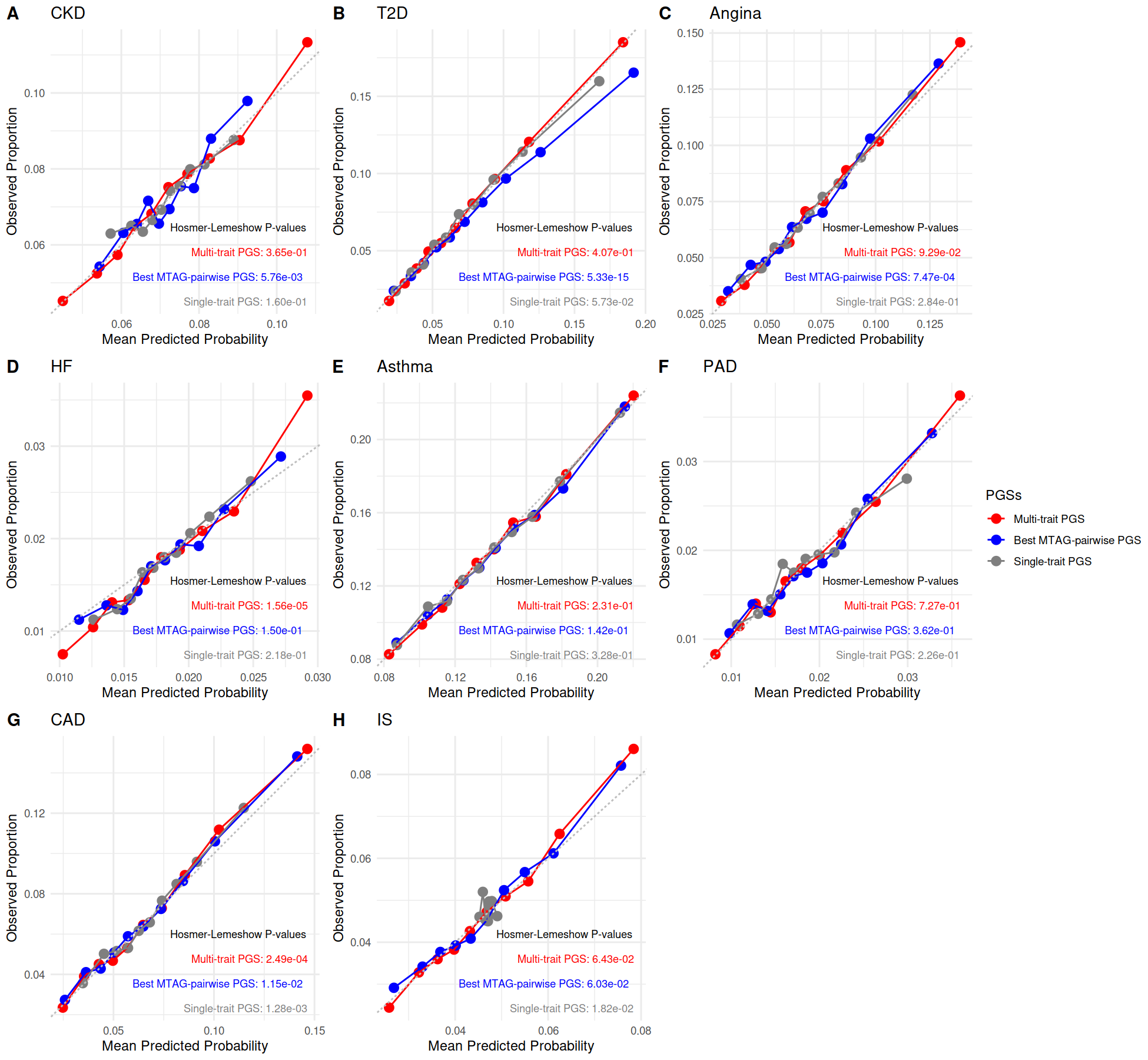

Calibration plots for disease risk prediction across three polygenic score models—single-trait PGS (gray), best MTAG-pairwise PGS (blue), and multi-trait PGS (red)—for eight diseases. Each plot compares observed and predicted disease probabilities across deciles of predicted risk. Hosmer-Lemeshow test P-values are provided to assess goodness of fit. Across all eight diseases, the multi-trait PGS models demonstrated strong calibration, with observed proportions closely aligned with predicted probabilities. In several cases, the multi-trait model outperformed the single-trait and MTAG-pairwise models in terms of calibration accuracy, as indicated by higher Hosmer-Lemeshow P-values.

**Supplemental Figure 2. Prevalence across PGS quantiles for eight diseases.**

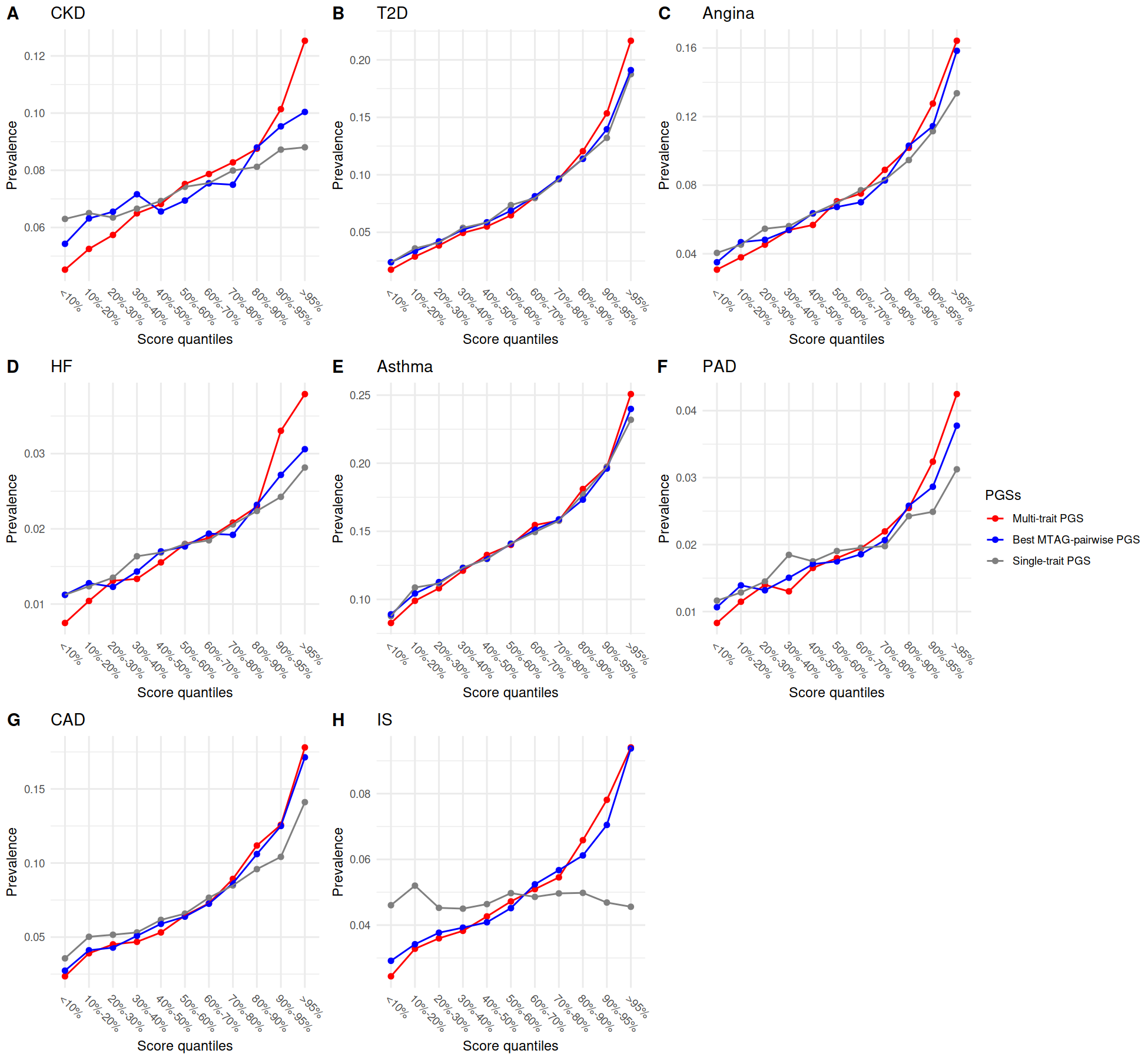

Prevalence of eight diseases across deciles of polygenic risk score distributions in the UK Biobank testing set, shown for three models: single-trait PGS (gray), best MTAG-pairwise PGS (blue), and multi-trait PGS (red). Across all diseases, the multi-trait PGS consistently demonstrated the highest disease prevalence in the top risk decile and the greatest separation between extreme quantiles, indicating improved stratification of individuals by genetic risk compared to single-trait and MTAG-pairwise models.

**Supplemental Figure 3. Calibration of multi-trait PGS models in internal and external validation datasets across eight diseases.**

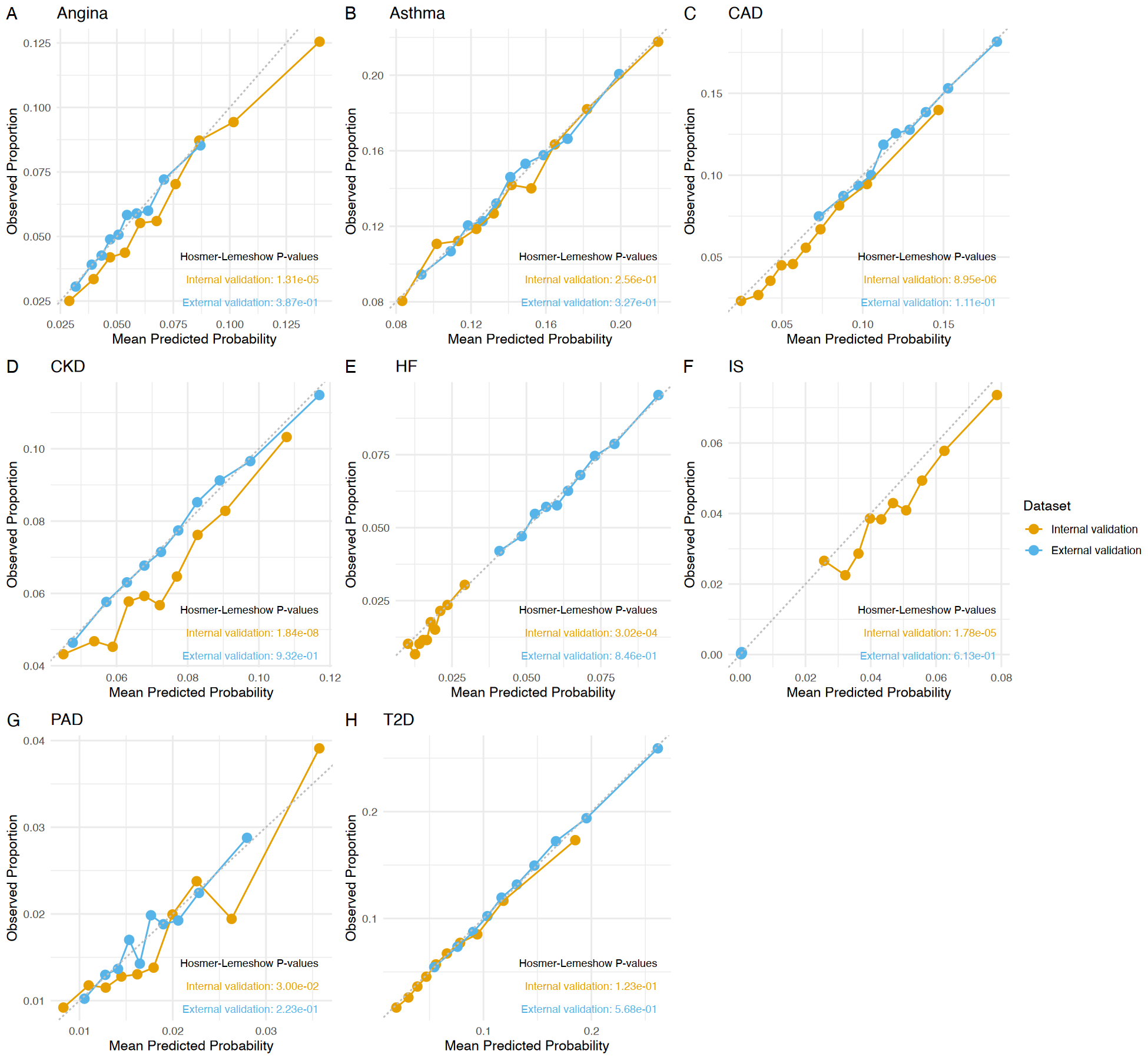

Calibration plots showing observed versus predicted disease probabilities for multi-trait PGS models in the internal validation cohort (non-British White UKB participants, orange) and the external validation cohort (European ancestry individuals from All of Us, blue). Each curve plots observed disease prevalence across deciles of predicted probability. Hosmer-Lemeshow P-values are provided for both cohorts to assess goodness of fit. Overall, the multi-trait PGS models demonstrated good calibration in both datasets, with slightly reduced calibration performance in the external validation cohort, likely reflecting cohort-level differences in SNP availability and phenotype distributions.

**Supplemental Figure 4. Prevalence across PGS quantiles in internal and external validation datasets for eight diseases.**

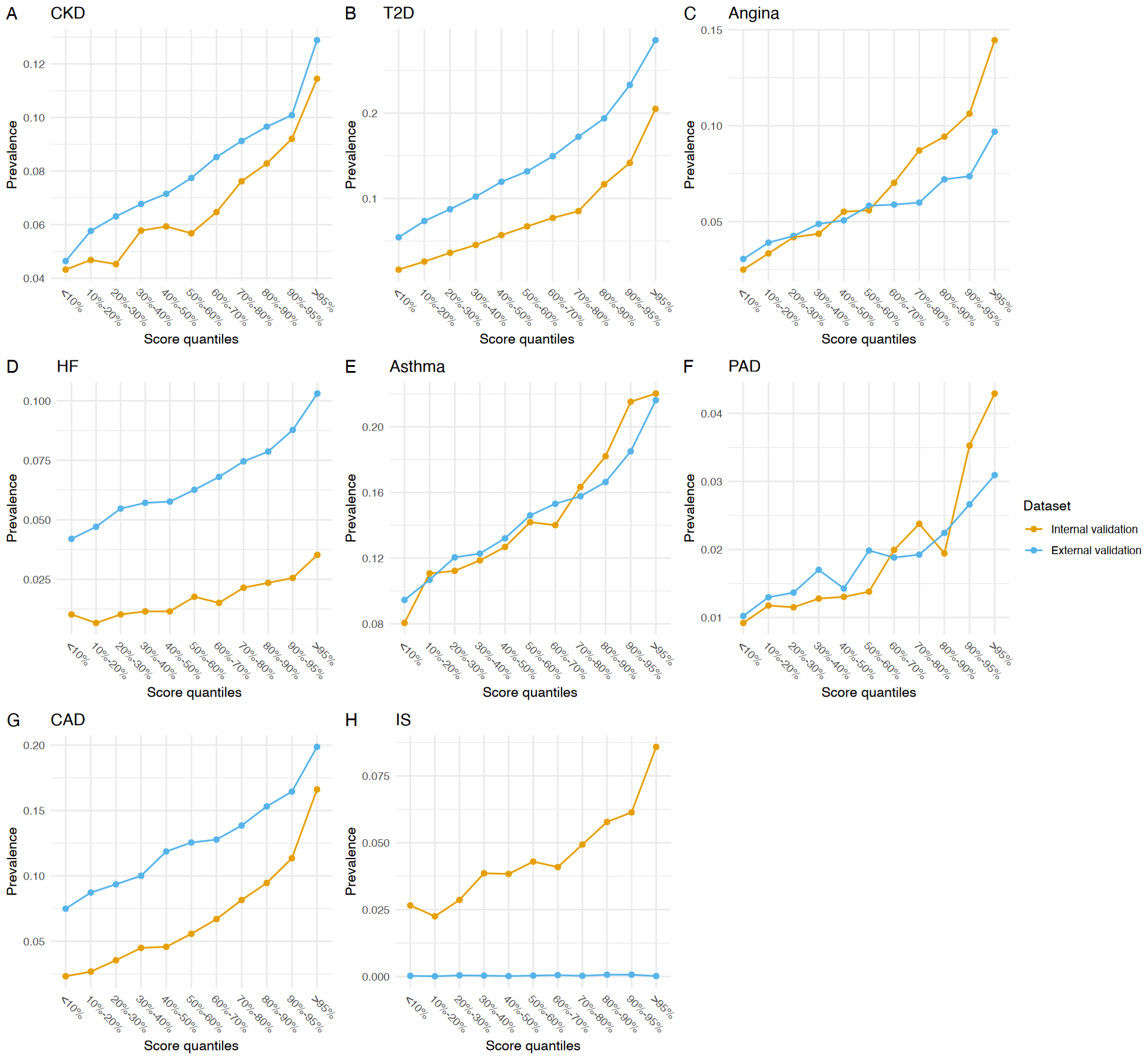

Prevalence of eight diseases across deciles of multi-trait polygenic score distributions in the internal validation dataset (non-British White UKB participants, orange) and the external validation dataset (European ancestry participants from All of Us, blue). In both cohorts, disease prevalence generally increased with PGS quantiles, reflecting the ability of multi-trait PGSs to stratify individuals by risk. However, patterns and effect magnitudes varied by disease and dataset. Notably, external validation showed consistently higher prevalence in the upper deciles for several diseases such as HF, whereas internal validation demonstrated sharper stratification for others, underscoring potential differences in cohort characteristics and phenotype definitions.

**Supplemental Figure 5. Genetic correlations between 51 GWAS summary statistics.**

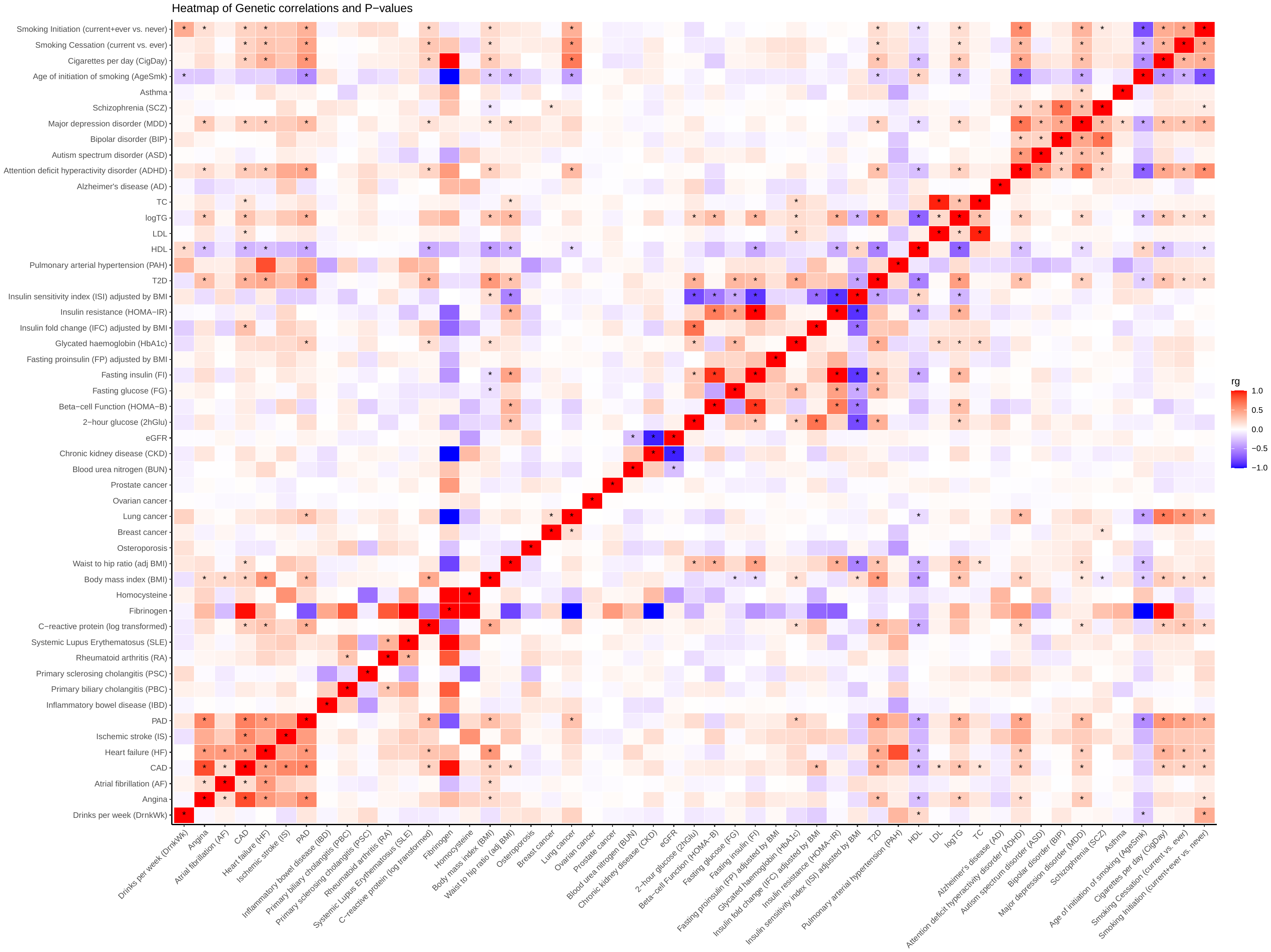
Heatmap displaying pairwise genetic correlations (rg) between 51 phenotypes used in the construction of multi-trait PGSs. Color intensity represents the magnitude and direction of the genetic correlation (red = positive, blue = negative), with statistically significant correlations highlighted by asterisks (based on Bonferroni-adjusted P-values). The results highlight widespread pleiotropy and cross-trait genetic sharing, particularly among cardiometabolic traits (e.g., T2D, BMI, WHR, CAD), neuropsychiatric traits (e.g., MDD, ADHD), and inflammation-related biomarkers (e.g., CRP, fibrinogen). These correlations underpin the rationale for multi-trait PGS integration to enhance prediction and capture comorbidity-related genetic risk.

**Supplemental Figure 6. Phenotypic associations with raw discordance scroes across eight diseases.**

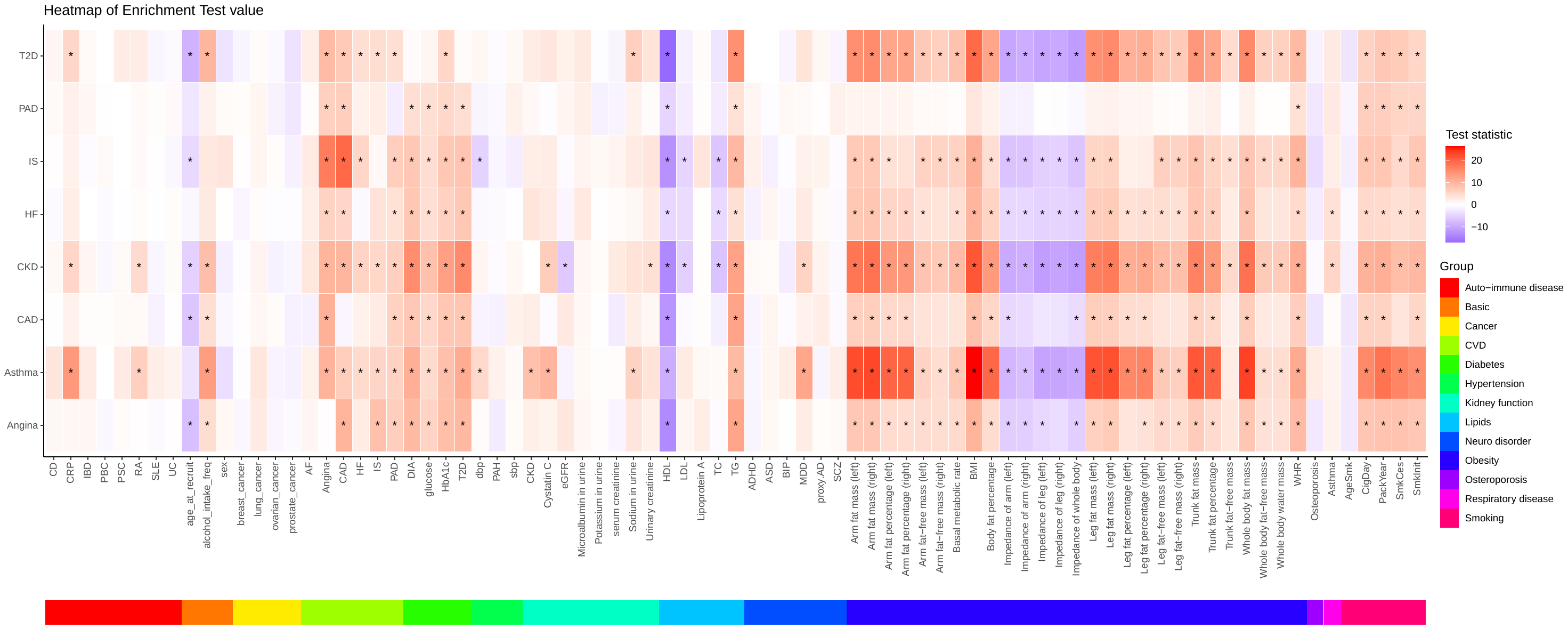

Heatmap showing standardized association statistics (z-scores) between the discordance score and 86 phenotypes among cases of eight diseases. The discordance score was defined as the difference between raw multi-trait PGS predictions and single-trait PGS predictions. Rows represent disease-specific analyses, and columns represent phenotypes grouped into 13 clinical or biological categories. Asterisks indicate phenotypes with statistically significant associations after Bonferroni correction. Notably, high discordance scores were associated with comorbid traits such as obesity, cardiovascular biomarkers, and neuropsychiatric disorders, supporting the hypothesis that multi-trait PGSs capture individuals with greater disease burden due to comorbidity. This analysis complements standardized results shown in Figure 4, supporting the robustness of associations between discordance scores and phenotype burden. Strong associations were particularly noted for metabolic and cardiovascular traits, reinforcing the utility of multi-trait PGSs in capturing comorbidity-driven genetic risk.

**Supplemental Figure 7. Calibration of prediction models integrating multi-trait PGSs and non-genetic risk factors.**

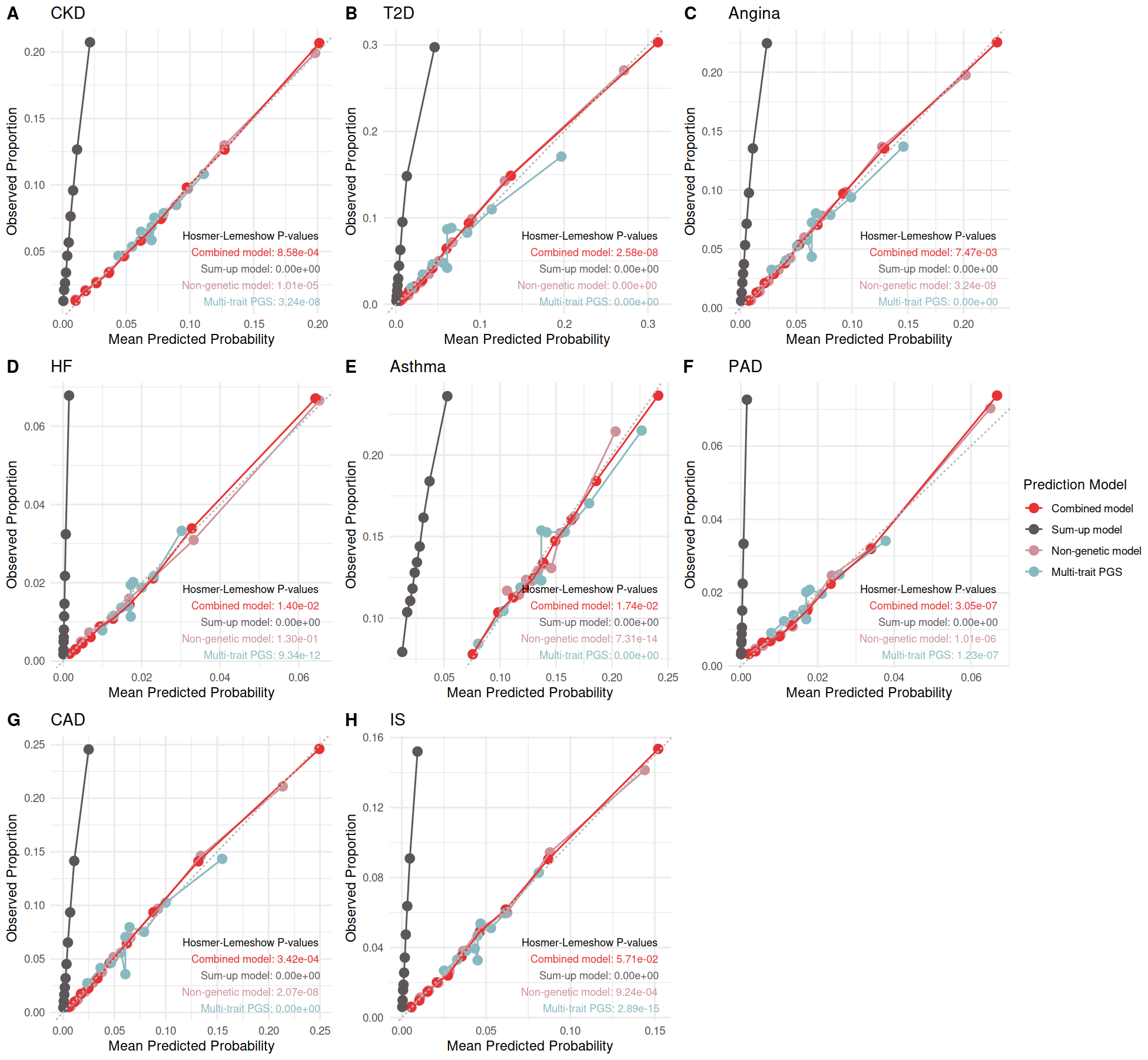

Calibration plots comparing observed and predicted disease probabilities across four risk prediction models: multi-trait PGS only (blue), non-genetic model (pink), sum-up model combining separate predictions (purple), and a combined model trained jointly on all predictors (red). Hosmer-Lemeshow P-values are provided for each model. Across most diseases, the combined model exhibited the best calibration, with predicted probabilities closely aligning with observed prevalence across risk deciles. The sum-up models tended to underperform, emphasizing the added value of integrating genetic and non-genetic information for accurate individual-level risk estimation.

**Supplemental Figure 8. Disease prevalence across score quantiles for prediction models integrating multi-trait PGS and non-genetic risk.**

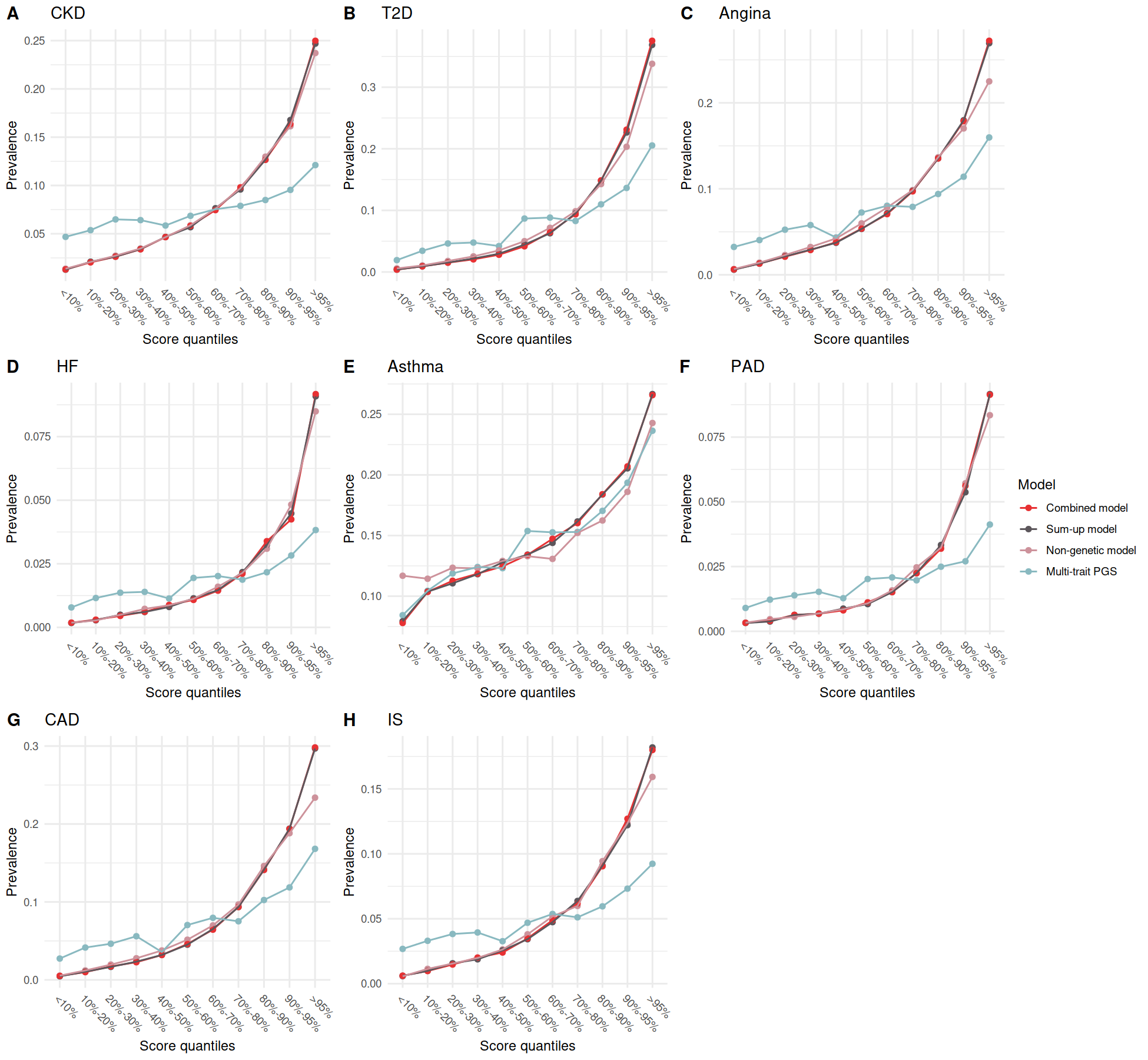

Prevalence of eight diseases across quantiles of predicted risk scores for four models: multi-trait PGS only (blue), non-genetic model (pink), sum-up model (purple), and combined model (red). Across all diseases, the combined and sum-up models exhibited the strongest stratification, with disease prevalence increasing most sharply across the score distribution. This trend was particularly pronounced in the top decile, highlighting the improved risk discrimination achieved by jointly modeling genetic and clinical predictors. The results demonstrate that integrating multi-trait PGSs with basic non-genetic risk factors substantially enhances population-level risk stratification.

**Supplemental Figure 9. Elbow plots for determining the number of genetic subgroups across eight diseases.**

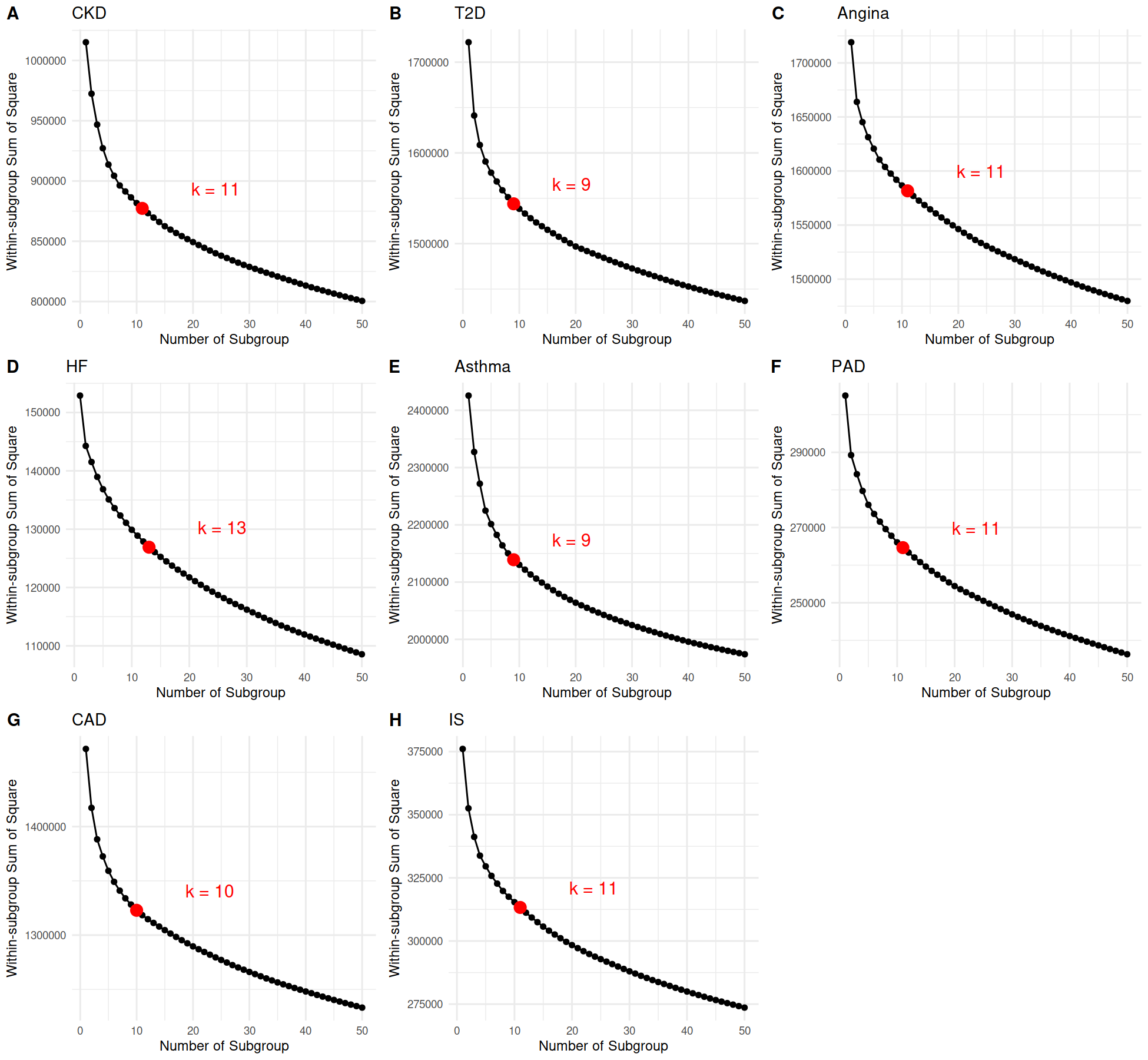

Elbow plots showing the within-cluster sum of squares (y-axis) versus the number of subgroups (x-axis) for hierarchical clustering of genetic profiles among disease cases. The optimal number of subgroups (k) for each disease was selected based on the inflection point ("elbow") in the curve, indicated by a red dot. The selected number of subgroups ranged from 9 (e.g., for T2D and asthma) to 13 (for HF), reflecting variation in the underlying genetic heterogeneity across diseases. These subgroup definitions were used for downstream analyses of genetic architecture and phenotypic variation.

**Supplemental Figure 10. Associations between phenotypes and genetic subgroups across seven diseases.**

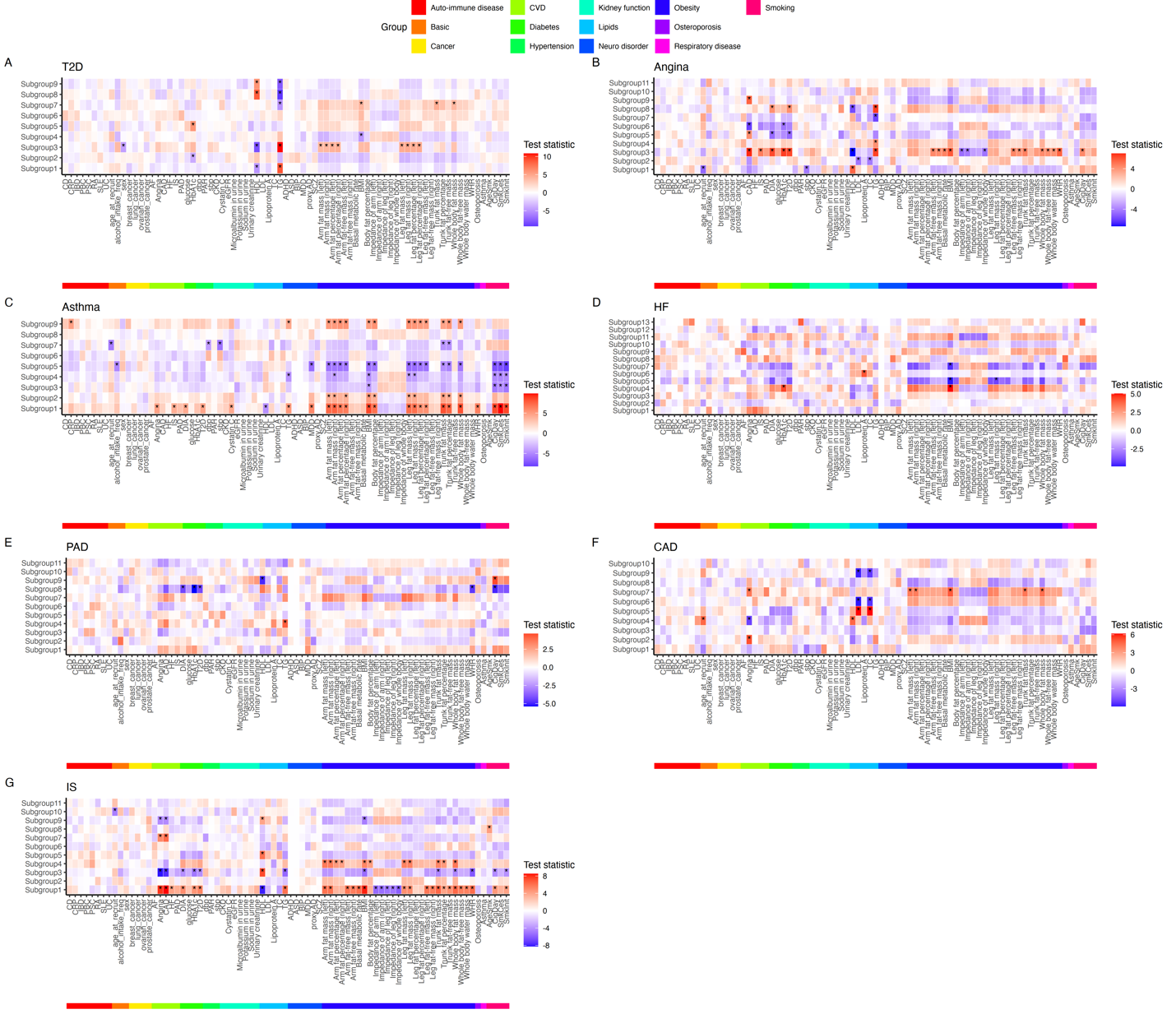

Heatmap showing statistical associations between genetic subgroups (rows) and 86 clinical and biomarker phenotypes (columns), grouped by biological category. Values represent test statistics (e.g., t-scores), with red indicating positive and blue indicating negative associations. Asterisks denote significant associations after Bonferroni correction. Subgroups exhibit distinct phenotypic profiles. Seven diseases were included for seven panels.
